# Disparate Impacts on Online Information Access during the COVID-19 Pandemic

**DOI:** 10.1101/2021.09.14.21263545

**Authors:** Jina Suh, Eric Horvitz, Ryen W. White, Tim Althoff

## Abstract

The COVID-19 pandemic has stimulated a staggering increase in online information access (*1, 2*), but the extent to which different communities of internet users enlist digital resources to meet everyday needs varies (*2-4*). We analyze 55 billion everyday web search interactions across 25,150 US ZIP codes and demonstrate that there were disparate impacts of the pandemic on online information access across several information domains, including health and pandemic-relevant online resources (e.g., online learning, online food delivery). Among many findings, we show that ZIP codes associated with higher proportions of Black residents intensified their access to unemployment resources, and ZIP codes associated with lower income reduced their access to health information resources relative to their counterpart ZIP codes. Because these disparate impacts on the access to online information may result in downstream offline gaps in health, education, employment, and well-being (3), public health interventions should target potential barriers to accessing the necessary digital resources and provide adequate support to meet the intensified digital resource needs.

**One Sentence Summary:** Large-scale web search logs reveal disparate impacts on online health, education, unemployment, and food information access.

## Introduction

Socioeconomic and environmental factors play a significant role in the health and well-being of individuals and communities (*5-7*). Despite pandemic-driven efforts to close the long-term and emergent health equity gap (*6*), studies during the COVID-19 pandemic have demonstrated that socioeconomically and environmentally disadvantaged subpopulations have been disproportionately and negatively affected by the disease (*8-10*), with threefold higher infection rates and sixfold higher death rates in predominantly black US counties than in white counties (*11*). In recent decades, digital access has also gained attention as an important factor modulating health outcomes, as individuals harness the internet to seek health information and to access healthcare services (i.e., telehealth, online pharmacy) (*4*). During the COVID-19 pandemic, digital engagement in resources across health, educational, economic, and social needs grew in importance because of lockdown mandates, social isolation, and economic burdens (*1, 2, 12*) as well as due to internet-based communication methods employed by public institutions, such as the online dissemination of COVID-related information by the World Health Organization (*2*).

Unfortunately, disparities in digital access also reflect socioeconomic and environmental dimensions of variation (*13*). The most basic form of digital inequality, the so-called first-level digital divide, manifests itself as the difference between adequate and inadequate digital infrastructure and devices (i.e., access to technology or the quality of access) (*14*). Digital inequalities also manifest themselves as the differences in the usage of digital technologies and skills relevant to the usage of digital technologies, the so-called second-level digital divide (*15, 16*). Most recently, the third-level digital divide has been conceptualized as the differential ability to translate the use of digital technologies into favorable outcomes, particularly in offline realms such as occupational pursuits, healthcare, and social networking (*17, 18*). For example, individuals with a low socioeconomic status (SES) have been shown to be slower than their higher-SES counterparts when it comes to using information and communication technologies (ICTs) during childhood and the entire life course. As a result, they may wind up with smaller social networks and limited employment opportunities (*3*). Furthermore, even after controlling for internet access, those from higher SES or higher digital literacy integrate digital resources into their lives and use the internet for more “capital-enhancing” activities that are likely to result in more upwards mobility in the offline world (*16, 18, 19*). Just as the social, economic, cultural, and personal offline resources can affect engagement in the corresponding digital fields, digital exclusion and the lack of engagement in digital resources can lead to negative offline consequences (*20*) across the range of downstream outcomes in the domains of health (*3, 4, 21*), education (*22*), and employment (*23, 24*). Therefore, it is important to observe digital behaviors across subpopulations and scrutinize the role of digital inequalities in our society. In addition, disadvantaged subpopulations are already at a higher risk of COVID-19 infection and mortality with heavier pandemic-induced socioeconomic burdens, such that it is critical to ensure that digital inequalities do not exacerbate the disparate impacts of the pandemic even further (*2*).

In this study, we harness the centrality of web search engines for online information access to conduct a retrospective observational study on how the pandemic may have shifted people’s engagement towards or away from digital resources and how such shifts may reflect offline societal disparities. Hence, our study focuses on quantifying the impact of the pandemic on how offline exclusion (e.g., lack of sufficient economic resources, lack of health insurance) impacts changes to existing digital exclusion (e.g., reduced participation in online banking or eHealth). This study extends prior work on pandemic-related disparities, many of which concern the epidemiological dynamics of the pandemic (*8-11*). Leveraging web search interactions enables us to model users’ search “interests” which are reflective of their underlying resource needs (*25-27*), including the use of critical digital resources such as online educational sites in response to school closures, online food delivery information in response to restaurant closures, online social interactions in response to physical distancing and travel restrictions, or online unemployment and economic assistance in response to economic instability during the pandemic. Given that the pandemic has impacted everyone’s web search behaviors across many different topic categories, however closely related to the pandemic itself (*1, 28*), our goal and key contribution is to identify differences across communities in their digital behavioral “responses” to the pandemic and to discover potential barriers and challenges in accessing critical resources on the web.

Prior work on understanding digital disparities has relied on costly surveys, interviews, or self-reports (*29-31*) that require direct engagement with the study population in order to prompt a recounting of their past behaviors rather than passively observing their actual behaviors. Datasets from specific service providers (e.g., Wikipedia (*32*), Zearn.org (*33, 34*)), domains (e.g., telehealth (*35*), eHealth (*21*)) or geographic areas (e.g., Northern California (*21*)) do not capture digital behaviors across a broad spectrum of human needs and subpopulations and at fine geo-temporal granularities. Macroeconomic measures, such as unemployment rates, do not capture potentially unmet needs or access barriers (e.g., confusion around unemployment benefits (*36-38*)).

Conversely, web search logs are routinely collected on a near real-time basis and at large scales, providing unique opportunities to unobtrusively examine digital behaviors across a wide range of topics, geographies, and subpopulations as well as highlighting potential barriers and changes to such engagement behaviors (*39*). In fact, web search logs have enabled studies of human behaviors across many different domains (*40-43*), times (*44-47*), locations (*48, 49*), and to make inferences about the future or to identify risk factors (*28, 50–53*). In the context of the COVID-19 pandemic, such data has stimulated a prolific range of research on physical (*28, 54*), psychological (*55, 56*), and socioeconomic (*57, 58*) well-being (*1*).

We contribute to this literature by analyzing 55 billion everyday web search interactions across multiple devices and 25,150 US ZIP codes during the COVID-19 pandemic. In our work, instead of focusing narrowly on a single topic, we aim to examine a spectrum of broader information domains to capture a holistic view of the impact of the pandemic (*59*). Our dataset includes anonymized search queries to the Bing search engine and subsequently clicked web site URLs from those queries. Each search interaction is classified into the categories of health, education, economic assistance, and food access that cover a broad range of critical resource needs (Supplementary Table S4). We link the search interactions from each United States ZIP code to their respective per-ZIP code census variables that broadly cover five social determinants of health (SDoH) categories defined by the US Department of Health (*60*): (1) Healthcare Access and Quality (through health insurance coverage), (2) Education Access and Quality (through education level), (3) Social and Community Context (through race/ethnicity), (4) Economic Stability (through income and unemployment rate), and (5) Neighborhood and Built Environment (through population density and internet access). We divide our dataset according these SDoH factors and compare the magnitude of change in search behaviors between two ZIP code groups during the pandemic, where larger observed difference in the magnitude of change in search behaviors could indicate that one group’s response to the pandemic is more significant than the other in the level of interest in online information (e.g., health, unemployment) or a in accessing online resources (e.g., online remote learning). For example, we split our ZIP codes into low and high income groups (below and above $55,000 median household income) and compare the magnitude of change in health condition information queries (Fig. 1a). To disentangle the confounding effects of SES and race/ethnicity on behaviors and health (*61*), we compare changes in search behaviors on matched pairs of ZIP codes that are highly similar across these potentially confounding factors (Methods). We isolate the relative changes in search behaviors that occur concurrently with the pandemic using difference-in-differences approach (*62*), adjusting for yearly and weekly seasonality and for pre-existing, pre-pandemic disparities in query volume (Fig. 1b-d, Methods). Thus, we measure the disparate impacts of the pandemic on the intensification or reduction of search behaviors between the two ZIP code groups delineated by their distribution in a single SDoH factor (Fig. 1e). Finally, we apply the same process across all SDoH factors (Fig. 1f, Methods).

**Figure 1:**
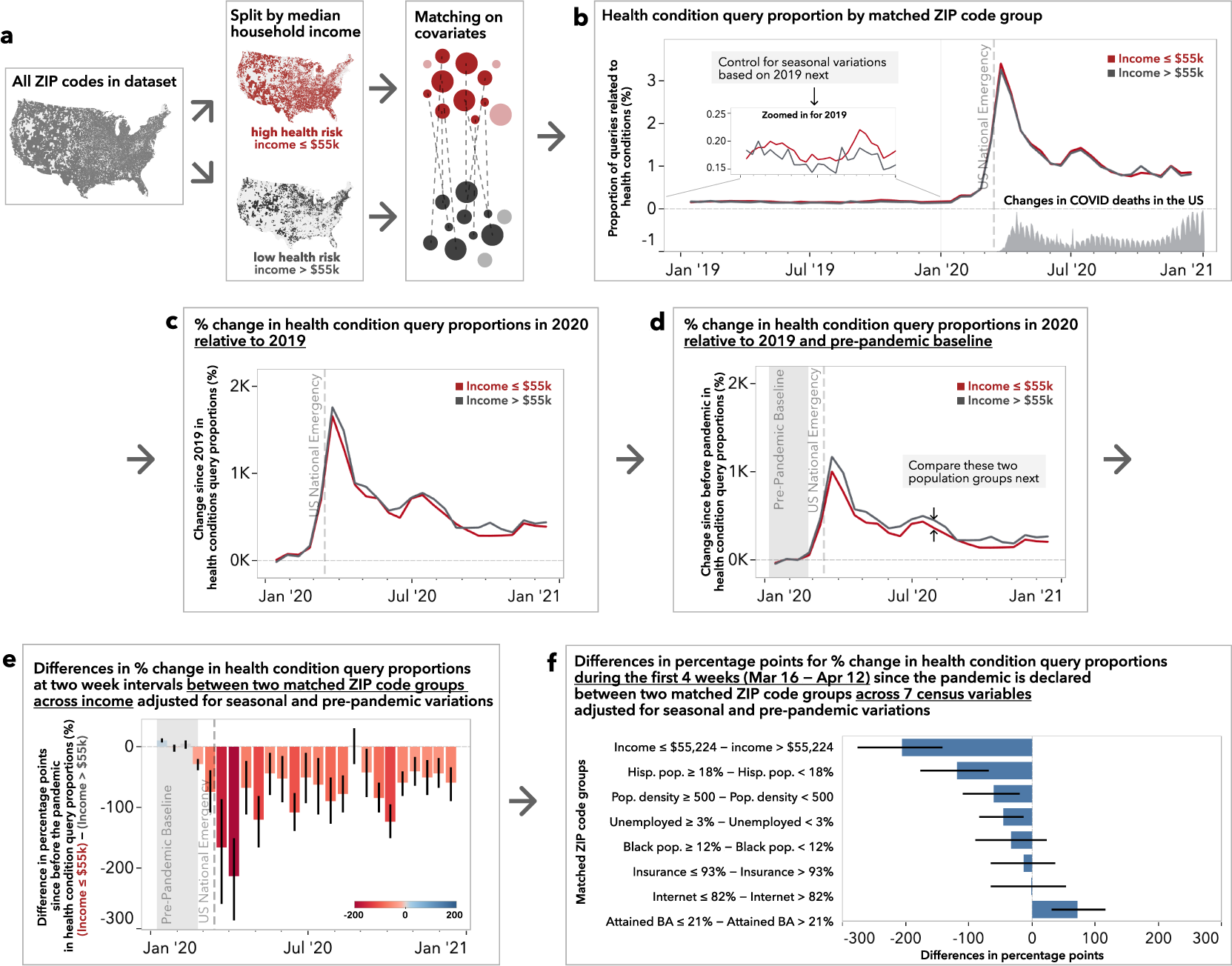
Quantifying disparities in online health information access. **a**, 25,150 ZIP codes above and below $55,224 median household income are matched to control for other confounding covariates (see Methods). **b**, The proportion of queries relating to a collection of health conditions in 2019 stay well below 0.25% of the total search queries across high income (gray) and low income (red) ZIP code groups, with mild seasonal highs around spring and fall and low income group exhibiting slightly higher health condition query proportion. This proportion increases dramatically to over 3% around the time the US national emergency was declared and is elevated throughout 2020 as COVID death rates change over time. After accounting for seasonal and weekly variations (relative to 2019, **c**) and the pre-pandemic baseline (relative to January 6 - February 23, 2020, shaded in gray in **d**), we isolate the percent change in health condition query proportions introduced during the pandemic where the differences between high- and low income groups start to emerge. **e**, We observe that low income ZIP codes experienced almost 200% less change in health condition queries compared to that of the high income groups right after the US national emergency is declared. **f**, When the same matching-based comparisons are performed across all SDoH factors during the first four weeks since the declaration of the pandemic in the US, ZIP code groups with lower incomes, higher proportions of Hispanic residents, higher population densities, and higher unemployment rates show significantly lower change proportions, while ZIP code groups with low education attainment show a significantly higher change in health condition query proportions. Error bars in all charts indicate 95% confidence intervals obtained through bootstrapping (N=500).

## Results

### Health information access

First, we examine the proportion of queries relating to a variety of health conditions (e.g., coronavirus and other health conditions including cancer or diabetes). Because the coronavirus, as the underlying cause of the pandemic, is at the forefront of everyone’s minds, the relative change in queries related to health conditions is almost 1000% higher than the pre-pandemic baseline. If all things were equal, we would see the same volume of response (i.e., the same relative change in query proportions) across all ZIP codes. However, given the higher rate of pre-existing health conditions, documented disparities in healthcare access, and higher COVID-19 case and mortality rates for low SES subpopulations (*8, 61*), we would expect to see that ZIP codes characterized by low SES would experience a greater intensification in their need for health information across a variety of health conditions and therefore increase their level of health information seeking behaviors more than their counterpart ZIP code groups. Instead, we find that ZIP codes associated with lower incomes show over a 200 percentage point smaller increase (95% CI [*−*287*, −*152]) in health condition queries than their higher income counterparts (Fig. 1e). This means that a ZIP code that was yielding a thousand health condition queries per month before the pandemic makes about ten thousand such queries per month during the pandemic, but a similar ZIP code would only yield about eight thousand such queries per month if that ZIP code had lower median household income. We find that ZIP codes with higher proportions of Hispanic residents, higher population densities, and higher unemployment rates also responded to the pandemic with lower relative change in their health condition queries during the first four weeks (Fig. 1f). While ZIP codes with high (i.e., above population-average) proportions of Black residents (*≥*12%) do not seem to be affected as much as those with high proportions of Hispanic residents during the first four weeks, their response is lower during the months of August to November (Supplementary Fig. S13g). On the other hand, we find that ZIP codes with low education (*≤*21.1% with bachelor’s degrees) make over 70 percentage points more (95% CI [31, 117]) health condition queries compared to ZIP codes with higher education (Fig. 1f).

Prior research has shown that SES and demographics correlated with online health information seeking behaviors, highlighting the digital divide in health information access (*63, 64*). This divide has serious consequences. Through effective online health information-seeking behaviors, individuals can potentially make better healthcare choices and enjoy better health and well-being as a result, thereby reducing health disparities (*3, 4, 63, 65*). Unfortunately, our results suggest that ZIP codes with low income or high diversity distributions did not ramp up their health information seeking behaviors to the same degree as ZIP codes with high income or low diversity distributions – a gap which may exacerbate health disparities down the line (*3,66*).

### Economic assistance access

When we examine unemployment-related search interactions, we find that relative changes in unemployment-related search queries (e.g., “eligible for unemployment benefits”, “jobless claims”) closely follow those of reported unemployment claims by the Bureau of Labor Statistics (Supplementary Fig. S11). However, the impact of the pandemic on ZIP codes with higher proportions of Black residents and their increase in unemployment search queries is almost three times the increase corresponding to ZIP codes with lower proportions of Black residents (Fig. 2a), with a 3026% increase in query proportions for ZIP codes with higher proportions of Black residents compared to an over 1365% increase for their counterparts, resulting in a 1,661 percentage point difference (95% CI [260, 2374]) (Fig. 2b). We find another surge in search queries that resulted in an over 1000% increase in the proportion of clicks on state-specific unemployment websites past July 2020, at which point the expanded federal supplement to unemployment insurance benefits expired (Fig. 2c). During the month of August, ZIP codes with higher proportions of Black and Hispanic residents present 789 (95% CI [595, 957]) and 716 (95% CI [351, 1043]) percentage points more in their change in clicks to unemployment sites, indicating that ZIP codes with higher proportions of Black and Hispanic residents may have required additional long-term unemployment benefits. Conversely, ZIP codes with lower educational attainment levels experienced 517 percentage points less (95% CI [*−*1009*, −*81]) in the change in state unemployment site visits (Fig. 2d).

**Figure 2:**
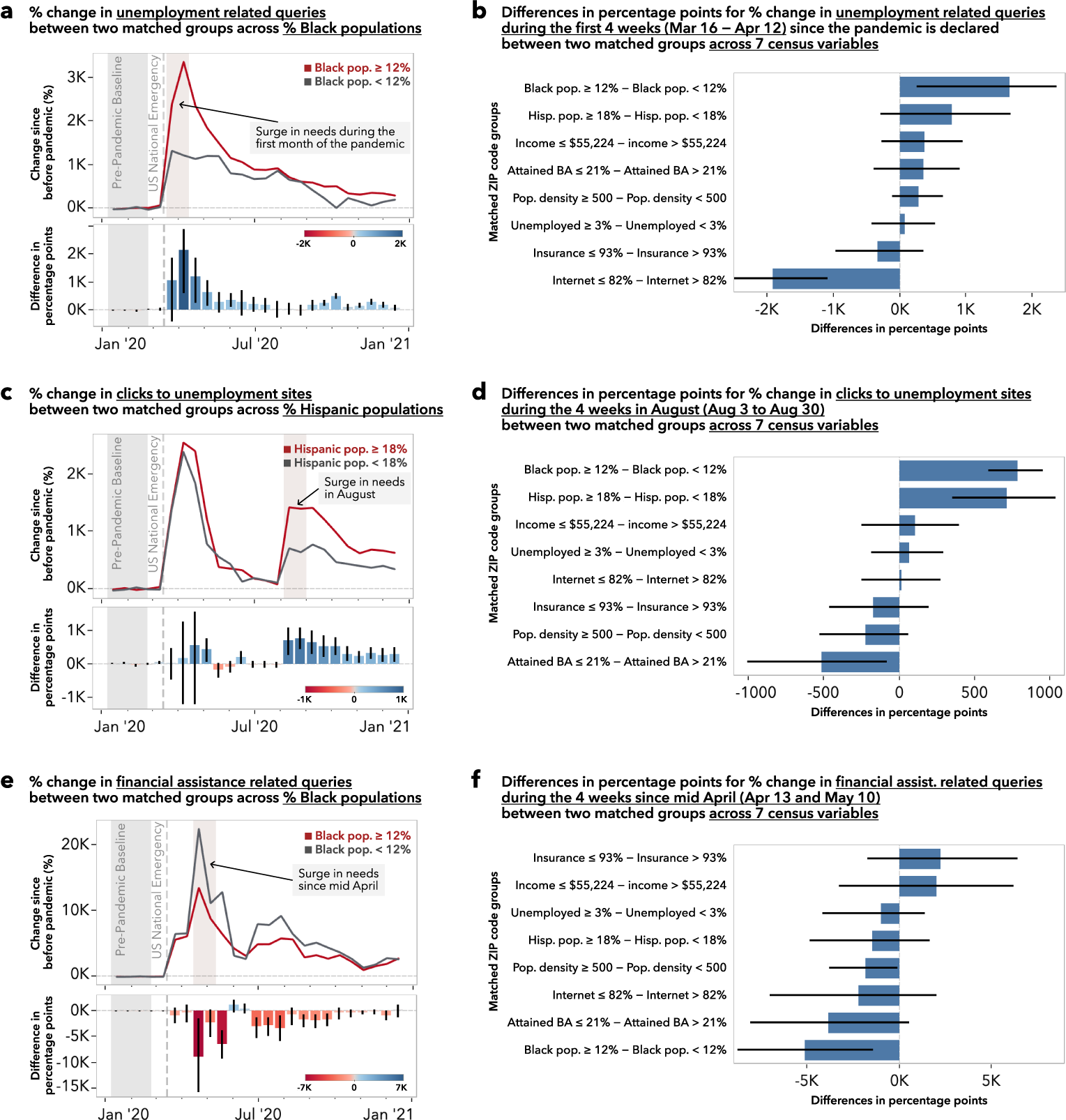
Disparities in online economic assistance access. **a**, The surge in unemployment related search queries peaks during the first month since the declaration of the pandemic and tapers off over the year 2020. During this first month, ZIP codes with higher proportions of Black residents (*≥* 12%) have expressed up to 3,358% more unemployment related queries while ZIP codes with lower proportions of Black residents (*<* 12%) have expressed 1,320% more. **b**, Across the seven census variables, ZIP codes with higher proportions of Black or Hispanic residents, and lower income populations experience greater changes in unemployment related queries during this first month. **c**, When we examine search queries that led to clicks in state unemployment sites, we see a second surge in August, with ZIP codes with higher proportions of Hispanic residents (*≥* 18%) experiencing more than double the change in clicks in state unemployment sites compared to ZIP codes with lower proportions of Hispanic residents (*<* 18%). **d**, We observe that ZIP codes with higher proportions of Black and Hispanic residents experience greater change in clicks in unemployment sites during the month of August, but ZIP codes with low educational attainment express less change in clicks in unemployment sites. **e**, Search queries related to financial stimulus were at their peak in late April, right after the time that the first stimulus checks were deposited on April 11. **f**, However, throughout the year and especially during the four weeks since mid-April, ZIP codes with higher proportions of Black residents experienced a smaller change in financial stimulus related queries than ZIP codes with lower proportio9ns of Black residents.

Internet access and digital engagement is an important form of human capital that allows efficient access to information, increases in economic opportunities, and ultimately leads to better prospects and economic stability (*67*). During economic hardships and especially during the pandemic, the internet can be an efficient way for governments and institutions to deliver interventions and can lower barriers to accessing economic assistance or welfare services (e.g., https://www.usa.gov/food-help provides a comprehensive list of resources for food assistance).

Unfortunately, the pandemic imposes multi-layered barriers to accessing crucial economic assistance because low SES subpopulations are more likely to suffer economically from the pandemic (*68*) and deprioritize improving digital access as a consequence (*3*). To understand the economic impact of the pandemic, we examine behaviors for accessing unemployment and financial assistance on the web.

Our results suggest that ZIP codes with lower educational attainment level, even while controlling for other factors such as the distribution of internet access, income, and race/ethnicity, are linked to barriers to accessing unemployment sites on the web. Since our difference-in-differences analysis already accounts for existing disparities, this difference in the increase in unemployment site visits signal a differential intensification of the demand in resources related to grappling with unemployment across ZIP code groups. Such “interest” in digital unemployment resources is not captured in reported claims that measure unemployment claims that are actually submitted, but can be readily observed in web search logs. The discrepancy between interests in unemployment benefits expressed online and officially submitted claims may suggest potential barriers in the successful submission of benefit applications (e.g., confusion, eligibility (*36, 37*)). Coupled with a low recipiency rate of unemployment benefits (*69*) and the association between unemployment accessibility and suicide risks (*70*), the mismatch between demands and claims are concerning.

April of 2020 was a prime occasion for financial assistance related queries (e.g., “loan forgiveness”, “stimulus check deposit”) because the first stimulus checks were deposited on April 11, 2020 (Fig. 2e). We find that financial assistance related queries increased by over 15,000% in mid-April on average, but ZIP codes with higher proportions of Black residents experience 5,119 percentage points less change (95% CI [*−*8809*, −*1407]) in financial assistance related queries between April 13 and May 10, 2020 (Fig. 2f). That means that if a ZIP code yielded 100 financial assistance related queries per month in mid-April of 2019, that ZIP code yields 16,700 such queries per month in mid-April during the pandemic, but only 11,600 queries for an otherwise similar ZIP code with a higher proportion of Black residents. Since we successfully controlled for other potential confounding factors such as income and education in our comparison, as shown in Supplementary Table S8, our result points to higher proportions of minority residents within ZIP codes, not necessarily the racial composition of the ZIP codes per se, as a plausible source for such disparity. Our finding highlights the need to further investigate potential barriers or causes that disproportionately prevent ZIP codes with higher proportions of Black residents from responding to pandemic-induced stimulus demands on the web.

### Shift to digital learning and food delivery resources

The COVID-19 pandemic brought a rapid and massive digital transformation to lives as mandated lockdowns forced people to transform and reimagine traditional interpersonal connections (e.g., going to school, getting food, or meeting friends) into virtual and digital ones. Unfortunately, digital inequalities worsen social and material deprivations and perpetuate existing disadvantages into a “digital vicious cycle” (*2, 72*). To understand the impact of the pandemic on reinforcing this vicious cycle, we investigate two types of digitally mediated activities which would be presumed to be particularly sensitive to pandemic-induced limitations on in-person access: online remote learning and online food delivery services.

Statewide mandates in the US required many schools to close in-person learning as early as March 16, 2020 (*73*), and school districts scrambled to implement remote learning alternatives. Many parents, students, and teachers turned to free online resources such as Khan Academy to fill the gaps temporarily or permanently (*74*). There were also reported disparities in access to technologies or live virtual learning as well as absenteeism that stymied low income students (*75*). When we examined search queries that result in visits to free online learning resources (e.g., coursera.org, khanacademy.org),

During the first four weeks of the pandemic, there was an overall increase in the proportion of queries that led to online learning sites compared to before (seen as a positive percent change in Supplementary Fig. S20). During this time, we found that ZIP codes with lower income and higher proportions of Hispanic residents exhibited only half to two-thirds of the increase (percentage point difference 95% CI [*−*227*, −*109] and [*−*202*, −*46] respectively) in those queries relative to their counterpart groups (Fig. 3a). If a ZIP code yielded 100 search-led clicks to online learning sites per month before the pandemic, that same ZIP code would yield 500 such clicks per month during the pandemic, but only 300 such clicks would be observed for a similar ZIP code with lower income or a higher proportion of Hispanic residents, even after controlling for internet access (Fig. 3b). ZIP codes with higher proportions of Black residents and higher population densities exhibit a similar trend. Even though these free online learning resources are designed to be accessible and flexible, helping students to go at their own pace, we find that ZIP codes with low income or high proportions of diverse residents did not leverage them at the same level as their counterpart ZIP code groups during the pandemic.

**Figure 3:**
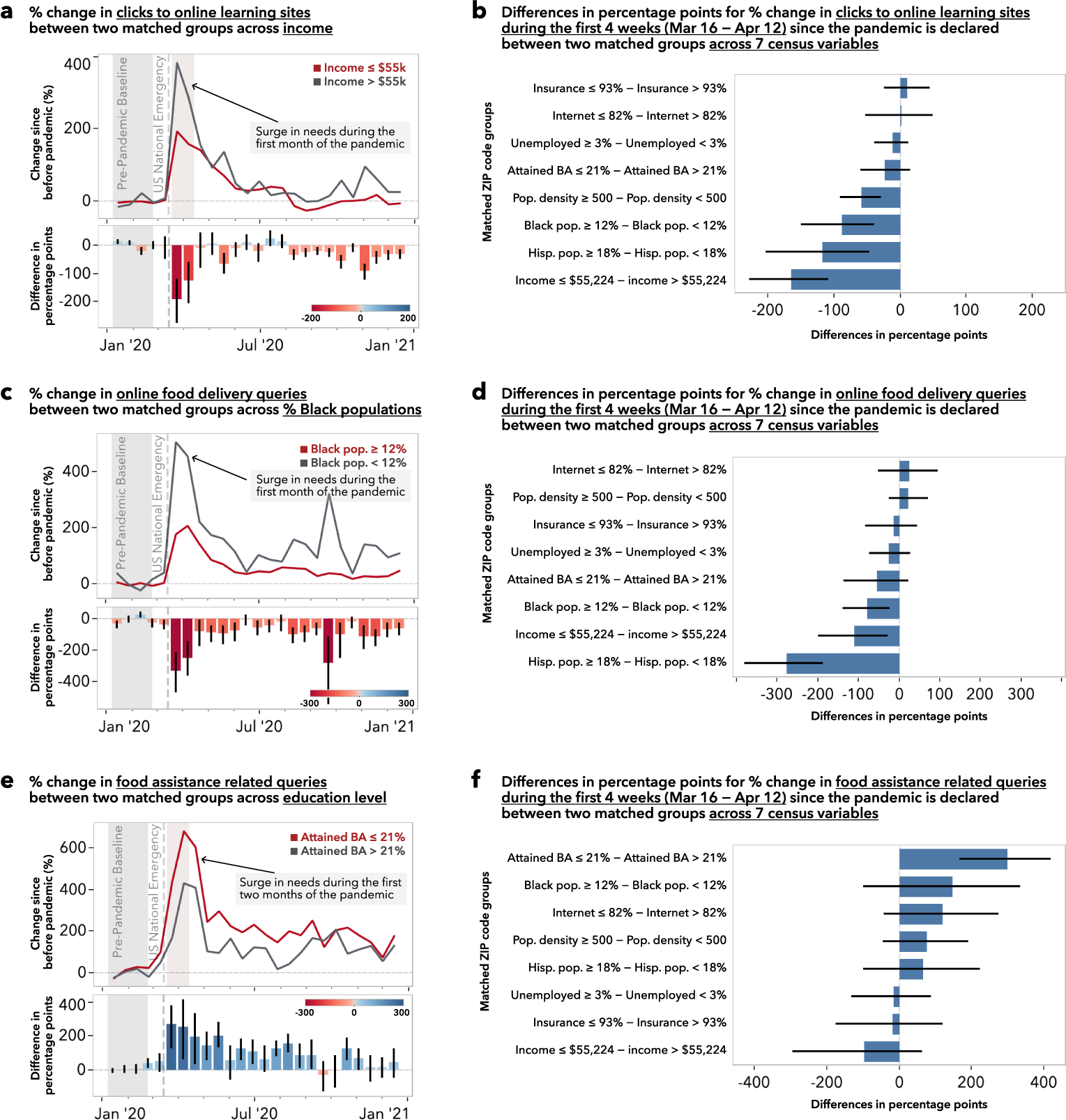
Disparities in shifting to digital resources. Engagement in critical online resources necessary during prolonged lockdowns or school/business closures were lower for ZIP codes with low income and high racial/ethnic diversity. **a**, Online learning sites played a significant role in filling in the gaps introduced by school closures at the beginning of the pandemic with an over 200% increase in engagement. **b**, However, ZIP codes with lower income and higher proportions of Black residents tend to access online learning resources less. In the new academic year (after August), while low income groups continued to show lower engagement, ZIP codes with higher proportions of Black residents show slightly higher engagement in online learning sites. **c**, With mandated restrictions on social gatherings, populations have transitioned to online-mediated social activities during the pandemic. **d**, For ZIP codes with a higher population density, where lockdown measures were more strictly enforced due to higher case and mortality rates (*71*), changes in online social activities search were higher. However, we see that ZIP codes with higher proportions of Hispanic residents show less change in online social engagement, even after controlling for population density or internet access, indicating potential racial/ethnic barrier or preference to accessing online social activities. **e**, Online food delivery services were also in high demand due to restaurant closures, with an over 250% increase at the beginning of the pandemic. **f**, However, ZIP codes with higher proportions of Black residents and poorly educated subpopulations showed a smaller change in online food delivery search throughout the pandemic, regardless of population density, income, or internet access.

On the other hand, during the fall academic period of 2020, the proportion of queries that led to online learning sites decreased compared to before (seen as a negative percent change in Supplementary Fig. S20). During this time, we found that ZIP codes with lower income and higher unemployment rates exhibited a smaller reduction (i.e., their change remained closer to the baseline, Supplementary Fig. S21e and h), but ZIP codes with a higher proportion of Black residents exhibited a larger reduction (Supplementary Fig. S21g).

In addition, school districts in low SES neighborhoods were more likely to be closed during the pandemic and less equipped to provide remote learning or at-home assignments, greatly reducing opportunities for both in-person and online learning for students with negative educational outcomes (*76, 77*). Our findings suggest that there exists unintended consequences of the public health policies that perpetuate a myriad of disadvantages, as education is such a crucial factor in digital literacy, socioeconomic status, and health.

COVID-19 fundamentally changed people’s purchasing and spending behaviors, as many of the restaurants, stores, and non-essential businesses were closed to in-person shopping (*78*). Spending on food delivery and groceries also increased significantly during the pandemic, with more people eating at home with a higher utilization of online e-commerce platforms for accessing food and groceries (*78,79*). When we examine search queries for online food delivery (e.g., “grocery delivery”, “deliver food”), we find that online food delivery queries increased by over 500% for ZIP codes with lower proportions of Black residents while those with higher proportions of Black residents only increased by over 170% (percentage point difference 95% CI [*−*382*, −*188], Fig. 3c,d). We found similar lessened engagement in online food delivery searches for ZIP codes with lower income and higher proportions of Hispanic residents (95% CI [*−*200*, −*29] and [*−*140*, −*24] respectively, Fig. 3d). These findings could be explained by the fact that low income subpopulations receive and seek more food assistance and tend to eat food away from home less frequently (*80*) and that such online food delivery services may not be accessible because they incur higher costs for consumers, given the markup and delivery surcharges.

ZIP codes with lower education attainment also experienced a 301 percentage point higher increase (95% CI [167, 419]) in queries for seeking food assistance (e.g., “Supplemental Nutrition Assistance Program“, “help with food stamps”, “free and reduced lunch”, Fig. 2e,f) relative to their highly educated counterparts. Unfortunately, those that relied on these traditional food assistance programs were left with severely limited choices during the pandemic because these programs do not extend to online purchase or delivery services (*81*). Our findings highlight a potential gap between the increased food assistance need, as illustrated by the increase in the online information seeking behavior *about* food assistance, and the ability to actually procure food goods through online food purchase and delivery services.

## Discussion

This study provides quantitative evidence that, across multiple domains during the pandemic, the relative intensification or reduction of online information seeking behaviors reflects the differences in socioeconomic and environmental characteristics of ZIP code groups. Prior studies have shown that access to digital resources and information and the incorporation of such digital technologies in everyday lives from childhood are crucial for upwards mobility (*3*). Although SES is an important factor in shaping disparities in digital access, prior research has shown that SES also impacts levels of web expertise and the utilization of digital resources for information-seeking activities (*19*). Low SES populations suffer from the lack of training and educational support key to building the necessary skills to make efficient use of digital access and tools (*13*), highlighting that simply making the internet more accessible may not level the playing field (*82*). In the context of the current COVID-19 pandemic, where digital access and resources became more critical due to prolonged at-home isolation and restrictions on in-person activities, communities and ZIP codes characterized by low SES may experience the compounding effects of multiple potential disadvantages that may manifest as disparate reactions to the pandemic in digital engagement. Our findings highlight information domains and communities with significant intensification and reduction in digital engagement due to the pandemic. We discover that ZIP codes with lower income or higher proportions of Hispanic residents are are less likely to mobilize web-based health resources, presenting similarly unfortunate consequences of eHealth initiatives that may disproportionately benefit digitally advantaged subpopulations (*3*). Additionally, the findings confirm that ZIP codes with lower income or high racial/ethnic diversity fell behind in the digital shift catalyzed by the pandemic (*2*), as these ZIP code groups did not leverage online learning or online food delivery resources as much as their high income and less diverse counterpart ZIP code groups.

We note the inherent limitations of studying digital engagement using digitally obtained data: This and other studies with online data can inadvertently exclude those who leave no or very little digital footprint (*3*). Our information sources provide signals about levels of activity, but we cannot study the details of changes in types of access if there is no engagement. Our analysis is also limited to the footprint of Bing as one of several search engines used for online information access, and Bing’s user population may not be fully representative of the United States population. Our study carefully controls for internet access, as measured by the census, such that any observed effects cannot be explained by differences in internet access across ZIP code groups. Our approach combines search log data with socioeconomic and environmental variables that are routinely captured through census tracks to examine the impact of such census variables on a diverse array of different topic areas of digital engagement at population scales. Our observed changes can only be attributed to ZIP code levels and not individuals because individual-level SDoH factors are not available and to preserve anonymity. Like any retrospective observational study, the potential for unobserved or uncontrolled variable confounding prevents us from making causal claims. However, we adjusted for the observed confounding through a matching-based and difference-in-difference based methodology (Methods). Our work provides a holistic characterization of digital engagement using broad categories spanning health, economics, education, and food, and we cannot make claims about specific subcomponents (e.g., individual keywords).

Our study is focused on studying the mediating impact of the pandemic on the impact of offline exclusion (i.e., SDoH factors) on digital exclusion (i.e., digital search behavior). Therefore, our analysis does not study the bidirectional nature of digital exclusion as theorized (*20*) and does not provide insights into the “third digital divide” (i.e., the differential offline outcomes that people obtain from their use of digital technologies (*18*)). This third dimension of digital divide is important in fully describing the negative reinforcement cycle resulting from the multilevel digital divide. Our current data cannot be directly used to discern whether different access behaviors are due to the lack of web expertise (i.e., digital literacy or search facility), the lack of awareness of the value of information (i.e., attitude towards information), or the lack of intangible resources like time and energy. However, concepts like digital literacy, which is an important factor in the embodiment of digital capital, can be quantified by careful examination of an individual’s search behavior. As prior research has shown, search interactions vary, based on the user’s familiarity with search engines or their domain expertise (*83, 84*). Quantifying digital literacy combined with a longitudinal observation of socioeconomic and environmental factors could provide empirical evidence for how digital literacy operates in the attainment of offline economic, cultural, and social capitals (*18*), and our large-scale, search-based methodology opens the doors of opportunities for monitoring such phenomena. In addition, we see value in follow-up, small-scale focused studies aimed at contextualizing individuals’ experiences of the crisis and measuring the effects of community-specific interventions (*2*). These community-specific interventions could include raising the level of digital literacy (e.g., education around web expertise or digital know-how) or improving the quality of digital access (e.g., high-speed, uninterrupted internet access or high-end equipment). Quality of access, especially through different device types or device specifications, has been highlighted as another important factor in recent digital divide research (*14*). While this study includes both desktop or mobile search interactions, more work is needed to understand the differential impact of the pandemic on desktop or mobile search interactions. These may also include non-digital methods because traditional methods (e.g., text messaging, handouts) have been shown to work better for low SES populations (*75*). Although the SDoH factors and outcomes reviewed in our analysis may not be the only variables of interest, our matching-based approach provides methodological robustness relative to traditional univariate analyses in observational studies by controlling for observed covariates (*85, 86*). Future research aimed at understanding digital disparities, therefore, must acknowledge the correlations between different SES, race/ethnicity, and social determinants of health (*87*) and leverage methods which embraces their interrelatedness (*88*).

This study presents a new approach to understanding digital disparities. It demonstrates that web search logs can be harnessed to characterize and deliver key insights about the disparate impact of global crises on the ways people utilize digital resources to meet everyday needs. Our observational study design is able to scale to a large population (billions of queries by millions of people) to quantify the disparities in digital engagement. Building on prior disparities research that advocated for a comprehensive look at SES factors including race/ethnicity (*61, 87*), our study emphasizes the inclusion of a broad set of factors and outcomes representative of the SDoH. Through the lens of SDoH factors, our findings highlight disadvantaged communities that may be struggling to overcome burdens induced by the pandemic and have disproportionately intensified or reduced their access to critical online resources. Therefore, future public health interventions should target both potential barriers to access that pull communities away from necessary digital resources as well as provide support to ensure that the intensified need for digital resources are adequately met.

## Materials and Methods

### Data Set and Study Population

Our source dataset consists of a random sample of 57 billion de-identified search interactions in the United States from the years 2019 and 2020 from Microsoft’s Bing search engine. Each search interaction includes the search query string, URLs of all subsequent clicks from the search result page, timestamp, and ZIP code from a reverse IP lookup. We excluded search interactions from ZIP codes with less than 100 queries per month so as to preserve anonymity. Our search dataset intentionally includes both desktop and mobile Bing search interactions in order to capture both search query sources. Although the quality of access, especially through different device types or device specifications, has been highlighted as another important factor in recent digital divide research (*14*), analysis on the differential search behaviors across device types is outside the study’s scope. All data were deidentified, aggregated to ZIP code levels or higher, and stored in a way to preserve the privacy of the users and in accordance to Bing’s Privacy Policy. Our study was approved by the Microsoft Research Institutional Review Board (IRB). Full details about the privacy and ethics review of the study can be found in the Ethics Declarations section.

While many Americans use other search engines such as Google, Bing’s query-based market share is estimated to be about 26.7% according to Comscore data (*89*). We focused on query-based metrics for estimating search market share because it captures end-users’ interaction with the search engine, including queries that may not have resulted in site visits^1^. To understand the validity of relying solely on Bing search data, we compared Bing and Google queries for matched categories longitudinally and found that the search trends are highly correlated (Pearson *r* = 0.86 to *r* = 0.98, Supplementary Figure S1).

One of our goals is to characterize the impact of socioeconomic and environmental factors on digital engagement outcomes. Unfortunately, data that combines individual-level search interactions with each individual’s socioeconomic and environmental characteristics at the US national scale does not exist, is difficult to capture, and invites privacy concerns. Instead, we use ZIP codes as our geographic unit of analysis. ZIP code level analysis can be limited because it cannot describe each individuals living in those ZIP codes. However, ZIP code level analysis can scale to nontrivial population sizes and has been repeatedly recognized and leveraged in population-scale and local/neighborhood-level research (*29, 90–95*). ZIP code level analysis also enables accounting for well-known issues associated with residential segregation and socioeconomic disparities (*13, 96*). We leveraged the available ZIP code level American Community Survey estimates using the Census Reporter API (*97*) in order to characterize the ZIP codes in our dataset.

Because of the multidimensional nature of socioeconomic status and its association to health outcomes, it is important to include relevant socioeconomic factors (*87*). We considered multiple socioeconomic factors including race, income, unemployment, insurance coverage, internet access, educational attainment level, population density, age, gender, Gini index, homeownership status, citizenship status, public transportation access, food stamp, and public assistance. We did not include some of the factors when they were highly similar to already included factors (e.g., % below poverty level is correlated to median household income, Pearson *r* = *−*0.624). In the end, we included eight census variables that represent all five categories of social determinants of health (SDoH) defined by the US Department of Health (*60*) to cover a broad range of socioeconomic and environmental factors. Under Healthcare Access and Quality, we included the percentage of the population with health insurance coverage (Table B27001). Under Education Access and Quality, we included the percentage of the population that attained a Bachelor’s degree or higher (Table B15002). Under Social and Community Context, we included the percentage of the population with Hispanic origin (Table B03003) and the percentage of the population with Black or African American alone (Table B02001). Under Economic Stability, we included the median household income (Table B19013) and the percentage of the civilian labor force that is unemployed (Table B23025). Under Neighborhood and Built Environment, we included the percentage of the population with a broadband or dial-up internet subscription (Table B28003) and the population density. We computed per ZIP code population density by joining area measurements from ZIP Code Tabulation Areas Gazetteer Files (*98*) and total population (Table B01003). We joined the search interaction data with the above SDoH factors on ZIP codes and excluded ZIP codes that did not have either search interactions or census data. The resulting 55 billion search interactions covered web search traffic from 25,150 ZIP codes in the US, and these ZIP codes represent 97.2% of the total US population. Supplementary Table S1 provides per-ZIP code summary statistics of our dataset.

### Disparate Impacts on Digital Engagement during the Pandemic

We leverage interactions with search engines to obtain signals about digital engagements where everyday needs are expressed or fulfilled through a digital medium, in our case Bing (*1*). In our study, we characterize digital engagement through modeling users’ search interests as expressions of underlying human needs (*1*), building upon prior work that uses search interactions to model “interests” which are either expressed explicitly through search queries or implicitly through clicks on results displayed on the search engine result page (SERP) (*25-27*). To gain a nuanced understanding of these search interactions, we categorize each search interaction into topics ranging from health access, economic stability, and education access using detectors on query strings and subsequently clicked URLs based on regular expressions and basic propositional logic. To capture the level of search interest in these categories in relation to all other categories of interest, we compute the proportion of total search queries that belongs to a specific category. For example, we compute the proportion of total search queries that contain health condition keywords such as cancer, diabetes or coronavirus to quantify the level of interest in engaging in health information seeking behaviors in relation to all other digital engagement behaviors. In another case, we examine search queries that result in subsequent clicks to state unemployment benefit sites to quantify the level of interest in unemployment benefits. Supplementary Table S4 enumerates the categories we examined with example query strings, URLs, and regular expressions.

Examining individual search keywords or subcategories has been pursued by others within and outside the scope of the pandemic. In our study, the use of broad categories spanning health, economics, education, and food is intended to capture a holistic view of the impact of the pandemic across many different needs (*59*). Accordingly, we do not make any claims about subcomponents within a category because studying the impacts of the pandemic on subcomponents is out of scope of this work.

Certainly, there exist search keywords that are more popularized by the current pandemic, such as “coronavirus” or “covid”, that also belong in the health information category. However, these keywords are not unique to the current pandemic and have existed before. As infrequent searches for “coronavirus” might seem in 2019, in our data, the query frequency of “coronavirus” in 2019 was similar to that of “mers” and certainly not zero (Supplementary Figure S2). In fact, the pandemic has had a large impact across many categories of interests (*1, 28*), not just for some that are highly relevant to the pandemic. For example, Suh et al. (*1*) has demonstrated that many of the ordinary search topics, such as “toilet paper”, “online games with friends”, or “wedding” were significantly impacted by the pandemic.

In addition, the focus on the level of interest through query proportions rather than query frequencies is helpful in our analysis. First, it helps with accounting for the baseline differences in search access between two populations. Second, this focus on relative measures of search query frequency helps adjust for changes in query volume over time, which is a common practice in Information Retrieval and web search log analysis (*99, 100*). Supplementary Figures S3-S10 illustrate the temporal variations in relative query frequencies (left) and in relative query proportions (right) in each query category for each of two matched groups across all SDoH factors. Adjusting for the baseline differences in search access allows us to remove the existing access differences between two groups, and the temporal trends of the query proportions between the two groups become much closer.

We have examined with rigor the degree to which the shifts in search interests were experienced differently by different ZIP code groups. Specifically, we estimate the differences across SDoH factors in the amount of changes in search interaction behaviors that are most likely introduced by the pandemic, by controlling for seasonal and non-seasonal query volume variations as well as for pre-existing, pre-pandemic between-group differences to isolate how a ZIP code group reacted to the pandemic during the period of March 16, 2020 to December 27, 2020. To do this, we first use a difference-in-differences method (*62*) to apply several corrections.

After we categorize each search interaction with our categories of interest, we count and aggregate them per time window (i.e., 2-week or 4-week intervals in our analysis) and per ZIP code (Fig. 1a). We compute the proportion of the total query volume represented by each category for these time windows to quantify the level of search interests in that category while removing undesired variations in the query volume over time (Fig. 1b). We denote the digital engagement at time *t* in category *c* as the fraction of the total number of queries at time *t*: *E*(*t, c*) = *N* (*t, c*)*/N* (*t*). From this, we control for yearly seasonal variations by subtracting the digital engagements of 2019 from that of 2020: *E*(*t*^2020^*, c*) *− E*(*t*^2019^*, c*). People tend to behave differently on weekends, and we observed a 7-day periodicity in our data, sometimes known as the “weekend effect” (*101*). Therefore, when comparing two years, it is important to account for the weekend effect. In order to highlight the actual differences that are not explained by weekend mismatches across years, we aligned the day of the week between both years (i.e., Monday, January 6, 2020 is aligned to Monday, January 7, 2019). In addition, we ensured that our comparison analysis included all seven days of the week (i.e., look at means across one or multiples of a full week) (Fig. 1c).

Finally, to compute the change in digital engagement during the pandemic since the time at which the US national emergency was declared on March 16, 2020, we subtract the query proportions between January 6, 2020 and February 23, 2020, a period we defined as the “pre-pandemic baseline” (Fig. 1d). Even though the national emergency was declared three weeks later, we use February 23, 2020 as the cut-off because individual states declared a state of emergency at different times between February 29 and March 15 of 2020 and to avoid partial weeks in our analysis. This process results in the change in digital engagement most likely attributable to the pandemic. Our estimate of the *relative change in digital engagement* in category *c* between before and during the pandemic is defined as:

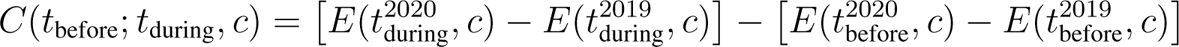

Or the relative *percentage change* in digital engagement *C*_perc_ is expressed as:

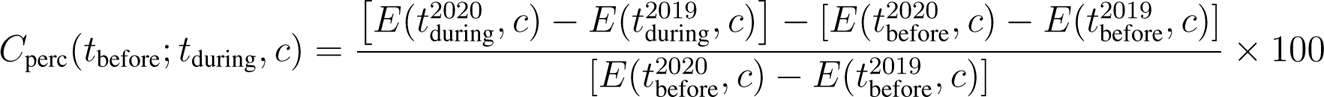

We acknowledge that there may exist a ZIP code with zero or very little search interactions for a given category, especially before the pandemic and in 2019. For example, “stimulus check” may only be relevant during the pandemic for certain ZIP codes. We cannot exclude these ZIP codes because we want a good representation and distribution of ZIP codes in our analysis. If a ZIP code makes only a handful of search queries on various health conditions, for example, but the number of queries increases dramatically due to concerns surrounding comorbidities and health complications, that is precisely the signal we hope to capture and observe across ZIP code groups. We mitigate this potential challenge of zero or near-zero baseline issues in several ways.

(1) Our regular expressions are inclusive of potential variations in expressing the categories, including expressions that are likely to occur before the pandemic and in 2019. (2) We aggregate search interactions in two or four-week windows, which consequently reduces the likelihood of having no or very little search interaction before the pandemic. (3) We also aggregate across thousands of ZIP codes that belong to a specific group (e.g., a group of ZIP codes with median household income greater than $55,224), where the likelihood of having no or very little search interaction before the pandemic for each group is 0%. (4) Instead of computing per-ZIP code difference-in-differences, we compute per-group difference-in-differences. In other words, we perform a within-group summation before taking the difference, which allows us to characterize the change in digital engagement for a “typical” ZIP code in the group.

Next, we aggregate these changes in digital engagements across two comparison ZIP code groups for each SDoH factor. For example, if we are examining the impact of having low income on the changes in digital engagement during the pandemic, we compare the average change in digital engagement of low income ZIP codes with the average change of the high income ZIP codes (Fig. 1d). Thus, we operationalize the disparate impacts of the pandemic attributable to a single ZIP code level SDoH factor by quantifying the differences in these changes in search behaviors between two ZIP code groups delineated by that factor (Fig. 1). In our analysis, we report the change in digital engagement as the percentages of the pre-pandemic baseline, *C*_perc_, where 0% denotes no change. We report the disparities in digital engagement between two comparison ZIP code groups as the percentage point difference where 0 denotes no difference (Fig. 1e,f). We formalize disparities in digital engagement in category *c* during the pandemic between high-risk ZIP code group *g*_high_ and low-risk ZIP code group *g*_low_ as:

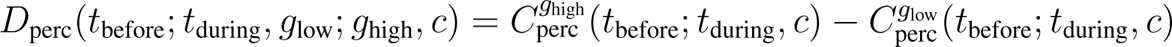

To obtain non-parametric confidence intervals, we conducted bootstrapping with replacement during this aggregation step (N=500). Supplementary Figures S12-S25 illustrate percent changes in each query category for each of two matched groups and their differences in percentage points across all SDoH factors.

### Matching to Control for Confounding

Our goal is to quantitatively estimate the independent impact of one socioeconomic factor on digital engagement while controlling for other factors to understand if and how a single factor independently impacts digital behaviors during a global crisis such as the COVID-19 pandemic. Specifically, we are interested in the impact of the eight SDoH factors: (1) median household incomes, (2) % unemployed, (3) % with health insurance, (4) % with Bachelor’s degree or higher degrees, (5) population density, (6) % Black residents, (7) % Hispanic residents, and (8) % with internet access. We use percent with internet access primarily to control the level of digital access within the ZIP code because internet access is a necessary precondition for web search. Many of the socioeconomic and racial variables are known to be correlated (*61, 87, 102*). This means that a univariate analysis of outcomes along one SDoH factor would likely be confounded by multiple other variables. In fact, within our dataset, we observed high correlation among many SDoH factors examined (Supplementary Table S3). For example, the median household income of the ZIP codes in our dataset is negatively correlated with the percentage of Black residents (Pearson *r* = *−*0.23) and is positively correlated with internet access (Pearson *r* = 0.66). Comparing high and low income groups without considering other factors would result in two groups of uneven distributions of race and internet access, among many other factors. Therefore, it is important to consider these factors jointly and adequately control for SES factors when analyzing outcome disparities (*61, 87*).

To disentangle the individual independent contributions of these SDoH factors when measuring disparities in online information access between groups, we employ a matching-based approach (*85, 86*), designed to create a comparable set of groups with similar covariate distributions. Because of the high degrees of spatial segregation in the US (*13, 96*), matching every ZIP code can be challenging. For example, for every ZIP code with low income and high proportions of Black residents, it is difficult to find a unique ZIP code with high income and high proportions of Black residents. Therefore, we perform one-to-one matching of ZIP codes with replacement and achieve better matches (i.e., lower bias). Theoretically, this is at the expense of higher variance, but given the size of our dataset, this downside was not a problem in practice. We use the MatchIt package (*103*) with the nearest neighbor method and Mahalanobis distance measure to perform the matching. We choose to perform matching on all covariates, instead of propensity scoring (*104*) which summarizes all of the covariates into one dimension. Importantly, we demonstrate in Section *Evaluating Quality of Matching Zip Codes* that this method leads to high-quality matches that are balanced across all covariates.

### Determining Treatment and Control Groups

For each of the SDoH factors, we first split all available ZIP codes into treatment and control groups using a threshold. We use a value close to the median to split the population into two groups for median household income ($55,224), % unemployed (3.0%), % with insurance (92.7%), % with internet access (81.8%), and % with Bachelor’s degree or higher (21.1%) because the mean and median of those factors across the ZIP codes are similar. In other cases, the distribution across the ZIP codes is highly skewed. For race/ethnicity, we use the rounded percentage of the national population for that race/ethnicity (12% for Black and 18% for Hispanic residents). For population density, we follow previous practices of urban-rural classification (500 people per square mile) (*105*). Supplementary Tables S1 and S2 outline descriptive statistics of our ZIP codes across SDoH factors as well as the national average and our chosen cutoff thresholds.

We consistently defined the treatment group as “high-risk” according to each of the dimensions of variation we specified (*66*). Therefore, our treatment groups are as follows: low income, high percentage of minority residents, low level of educational attainment, high unemployment rate, low insurance rate, low level of internet access, and high population density. For example, for income, we split the ZIP codes into a high-income group (median household income *>* $55,224) and a low-income group (median household income *≤* $55,224), where the low-income group is the treatment group. Then, for each treatment ZIP code, we look for a control (i.e., “low-risk”) ZIP code that closely matches it on all other SDoH factors (i.e., *|SMD| <* 0.25 to generate a matching pair of ZIP codes). We performed this matching on all ZIP codes, and we discarded ZIP codes for which we cannot find a good match. As demonstrated in Supplementary Table S6, this process retains at least 99.8% of the treatment ZIP codes in our matching process and the discarding of ZIP codes is a rare exception.

### Evaluating Quality of Matching Zip Codes

To gauge whether two ZIP code groups are similar across the SDoH factors and to determine the quality of matching while minimizing the potential confounding effects of these factors, we leverage Standardized Mean Difference (SMD) across ZIP code groups as our measure of comparative quality. The SMD is used to quantify the degree to which two groups are different and is computed by the difference in means of a variable across two groups divided by the standard deviation of the one group (often, the treated group) (*86, 106, 107*). In our analysis, we use *|SMD| <* 0.25 across all our SDoH factors as a criterion to determine that the two groups are comparable, following common practice (*85, 107*). For example, when we split our ZIP codes in half along median household income to create a high-income ZIP code group (median household income *>* $55,224) and a low-income ZIP code group (median household income *≤*$55,224) and examine the SMD of other SDoH factors, we find that all SDoH factors except % Hispanic residents and population density fail to achieve the necessary matching criteria of *|SMD| <* 0.25 prior to matching. This means that low-income ZIP codes are more likely to have less internet access, lower educational attainment level, less health insurance, more unemployment, and higher proportions of Black residents. We perform this evaluation process for all comparison groups to find that correlations among all SDoH factors pose threats to validity in univariate analyses. Supplementary Table S5 summarizes the mean SMD if we were to directly compare two ZIP code groups created by splitting the ZIP codes along the chosen split boundaries. Instead of such direct comparison, we perform matching and tune the caliper of the matching algorithm to determine a good match and to meet the *|SMD| <* 0.25 criterion between the two comparison groups across all covariates. Supplementary Table S6 summarizes the result of the matching operation with the maximum *|SMD|* being below 0.25, that is ensuring comparability across all covariates, between two ZIP code groups along all SDoH factors. Supplementary Tables S7-S22 enumerate pre- and post-matching balance assessments between groups for each SDoH factor.

### Estimating the Effect Size

After identifying treatment and control ZIP code groups with comparable distributions along all SDoH factors, we compare the outcomes (i.e., constructs of digital engagement such as online access to health condition information) between the matched ZIP code groups. This matching process estimates the Average Treatment Effect on the Treated (ATT), or specifically the effects of having low income on digital engagement while removing plausible contributions from all other observed factors. Due to the segregation and inequalities revealed by these factors, estimating the Average Treatment Effect (ATE) is practically impossible. One may opt to compute a local average treatment effect (LATE) and discard a large fraction of the U.S. population. However, such local estimates are easily misleading when the underlying population is not well understood and they fail to capture key populations in our study (e.g., low income and high proportions of Black residents). The ATT estimates in this study provide actionable insights on the effects of being at high risk (e.g., low income, low education, high racial/ethnic diversity) that can be used to suggest interventions to mitigate or reduce risk.

It is important to note that our matching process only partially incorporates what Helsper calls the digital impact mediators of access, skills, and attitudes (*20*). First, where digital access is concerned, though all search queries in the study presume some form of internet access, we do sample ZIP codes with varying levels of aggregate internet access, allowing us to control to some extent for internet access at the population level. Where digital skills are concerned, we do not incorporate direct measures of such technical or operational skills at either the individual or aggregate level, but we do incorporate measures of educational attainment such that we can partially control for this factor in our analysis. Finally, we do not control for individual-level or aggregate-level variation in attitudinal impact mediators such as self-efficacy, as that would be outside the scope of the study. Additional more detailed data would have to be collected and analyzed in order to fully disentangle the impacts of the SDoH factors under study here from such digital impact mediators.

## Data Availability

Raw US census data are publicly available through the Census Reporter API (https://censusreporter.org/). Geographical area measurements are available through the US Census Bureau (https://www.census.gov/geographies/reference-files/2010/geo/state-area.html). Seasonally adjusted US unemployment claims data for 2020 is available through the US Department of Labor (https://oui.doleta.gov/unemploy/claims.asp). The Bing data that support the findings of this study are available on request from the corresponding author with a clear justification and a license agreement. The Bing data are not publicly available.

https://www.census.gov/geographies/reference-files/2010/geo/state-area.html

https://oui.doleta.gov/unemploy/claims.asp

## Acknowledgments

We thank E. Pierson, E. Kiciman, the University of Washington Behavioral Data Science Group, Microsoft Research Human Understanding and Empathy Group, participants at seminars and talks for their support and comments.

## Author contributions

J.S., E.H., R.W., and T.A. were involved with the conceptualization of the study, and contributed to the design and refinement of the methodology. J.S. conducted data collection and analysis. All authors interpreted the data, drafted the manuscript, and critically contributed to the important intellectual content of the manuscript.

## Data and materials availability

Raw US census data are publicly available through the Census Reporter API (https://censusreporter.org/). Geographical area measurements are available through the US Census Bureau (https://www.census.gov/geographies/reference-files/ 2010/geo/state-area.html). Seasonally adjusted US unemployment claims data for 2020 is available through the US Department of Labor (https://oui.doleta.gov/unemploy/claims.asp). The Bing data that support the findings of this study are available on request from the corresponding author with a clear justification and a license agreement. The Bing data are not publicly available. Source code used for processing and analysis of the data is available on request from the corresponding author.

## Ethics declarations

The study (protocol ID 632) was reviewed by the Microsoft Research Institutional Review Board (OHRP IORG #0008066, IRB #IRB00009672) prior to the research activities. Microsoft Research is an industry-based research institution with a United States Department of Health & Human Services (HHS) federally registered IRB. The authors and the Microsoft Research IRB recognize the sensitive nature of the use of data collected from Microsoft users for research purposes. Our study followed the privacy and security regulations governed by Microsoft’s privacy statement as well as the federal ethical guidelines set forth by the HHS. All search data have been de-identified and aggregated prior to receipt by our study team such that no identifiable information was processed or analyzed. Microsoft Research IRB formally approved our study as “Not Human Subjects Research” to indicate that the activities do constitute research, but where the definitions of “human subject” and “identifiable private information” do not apply (as defined by 45*§*46.102(e)). Microsoft Research IRB certifies that our Human Subjects Review process follows the applicable regulations set forth by the Department of Health and Human Services: Title 45, Part 46 of the Code of Federal Regulations (45 CFR 46) (the Common Rule), and our Ethics Program promotes the principles of the Belmont Report in our research institution.

## Supplementary Materials

### List of Supplementary Materials

Table S1 to S22 Figures S1 to S25

**Table S1:** Means and medians of eight SDoH factors across 25,801 ZIP codes in our dataset. Population density is computed from the total U.S. population (*97*) divided by the total U.S. land area (*108*).

| SDoH Factor | ZIP Code Mean | ZIP Code Median | U.S. Mean | ACS Table |
| --- | --- | --- | --- | --- |
| Median Household Income | \$60,819.849 | \$55,224 | \$65,712 | B19013 |
| % Unemployed | 3.4% | 3.0% | 2.9% | B23025 |
| % Black or African American Alone | 8.3% | 1.6% | 12.8% | B02001 |
| % Hispanic or Latino | 10.1% | 4.0% | 18.4% | B03003 |
| % with Bachelor or Higher Degree | 25.9% | 21.1% | 33.1% | B15002 |
| Population Density (per sq mile) | 1573.966 | 108.421 | 92.830* | B01003 |
| % with Health Insurance Coverage | 91.1% | 92.7% | 98.4% | B27001 |
| % with Internet Access | 80.5% | 81.8% | 85.8% | B28003 |

**Table S2:** Split boundaries used to generate high-risk (treated) and low-risk (control) comparison groups based on ZIP code medians and U.S. national means of eight SDoH factors. Values that informed the choice of the split boundaries are bolded. For population density, we follow previous practices of urban-rural classification (*105*).

| SDoH Factor | ZIP Code Median | U.S. Mean | Boundary | Treated | Control |
| --- | --- | --- | --- | --- | --- |
| Median Household Income | **\$55,224** | \$65,712 | \$55,224 | $\leq \$55,224$ | $> \$55,224$ |
| % Unemployed | **3.0%** | 2.9% | 3.0% | $\leq 3.0\%$ | $> 3.0\%$ |
| % Black or African American Alone | 1.6% | **12.8%** | 12% | $\geq 12\%$ | $< 12\%$ |
| % Hispanic or Latino | 4.0% | **18.4%** | 18% | $\geq 18\%$ | $< 18\%$ |
| % with Bachelor or Higher Degree | **21.1%** | 33.1% | 21.1% | $\leq 21.1\%$ | $> 21.1\%$ |
| Population Density (per sq mile) | 108.421 | 92.830 | 500 | $\geq 500$ | $< 500$ |
| % with Health Insurance Coverage | **92.7%** | 98.4% | 92.7% | $\leq 92.7\%$ | $> 92.7\%$ |
| % with Internet Access | **81.8%** | 85.8% | 81.8% | $\leq 81.8\%$ | $> 81.8\%$ |

**Table S3:** Pearson correlation coefficients between SDoH factors across 25,801 ZIP codes in our dataset. These correlations reflect socioeconomic segregration and are controlled for through a matching-based approach (Methods). (*^∗^* = attained Bachelor’s degree or higher)

| SDoH factor | % Unemp. | % Black | % Hispanic | % BA* | Pop. Density | % Insurance | % Internet |
| --- | --- | --- | --- | --- | --- | --- | --- |
| M. H. Income | -0.25 | -0.23 | -0.07 | 0.74 | 0.10 | 0.41 | 0.66 |
| % Unemp. |  | 0.32 | 0.15 | -0.18 | 0.09 | -0.23 | -0.20 |
| % Black |  |  | 0.03 | -0.09 | 0.15 | -0.19 | -0.23 |
| % Hispanic |  |  |  | -0.07 | 0.21 | -0.35 | -0.04 |
| % BA* |  |  |  |  | 0.23 | 0.36 | 0.62 |
| Pop. Density |  |  |  |  |  | 0.00 | 0.10 |
| % Insurance |  |  |  |  |  |  | 0.41 |

**Table S4:**
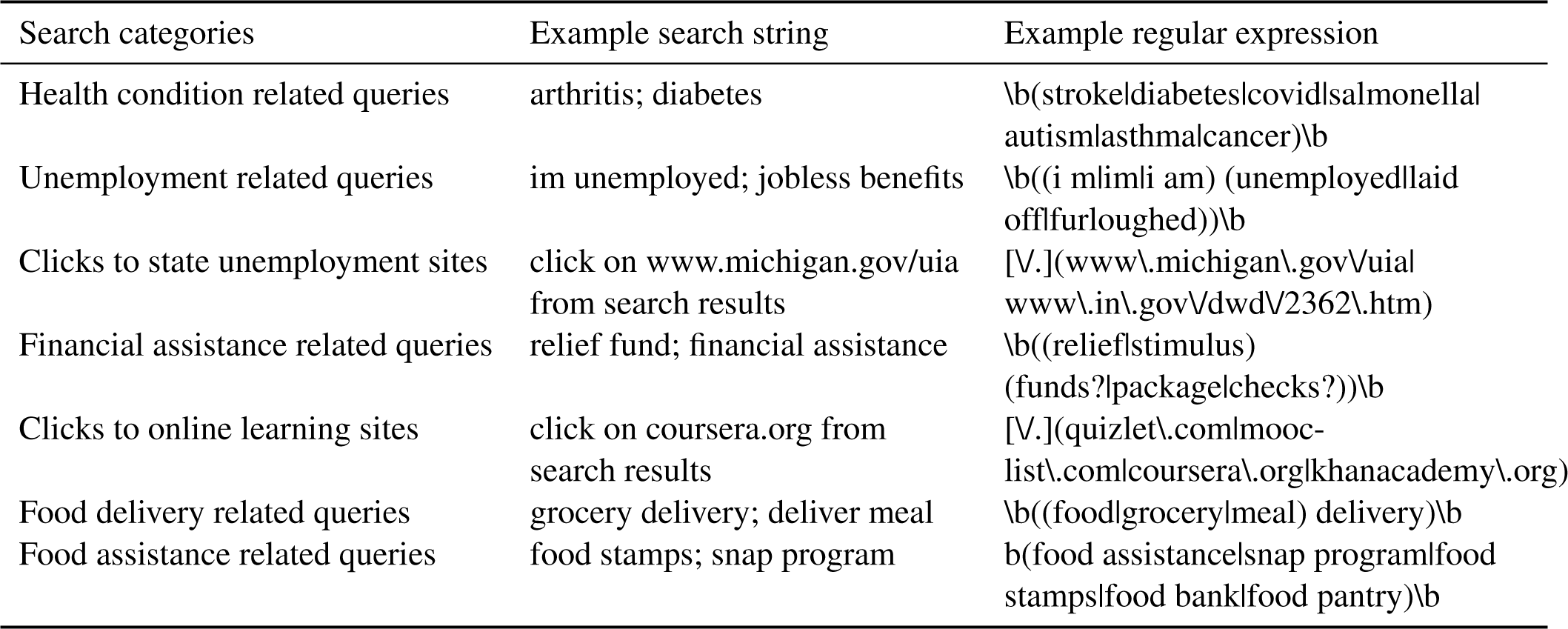
Seven search categories examined and their example search strings and regular expressions.

**Table S5:** Summary of *pre-matching* balance assessment and sample sizes across SDoH factors. *|SMD| >* 0.25 indicates that the two ZIP code groups created along the split boundary are different and, thus, not comparable.

| SDoH Factor | Split Boundary | Treated Count | Control Count | Max SMD | Mean SMD |
| --- | --- | --- | --- | --- | --- |
| Median Household Income | \$55,224 | 12637 | 12637 | 1.018 | 0.459 |
| % Unemployed | 3.0% | 12637 | 12637 | 0.385 | 0.269 |
| % Black or Afr. Am. Alone | 12% | 4945 | 20329 | 0.467 | 0.300 |
| % Hispanic or Latino | 18% | 4028 | 21246 | 0.591 | 0.218 |
| % with Bachelor or Higher | 21.1% | 12637 | 12637 | 0.975 | 0.416 |
| Population Density (per mi<sup>2</sup>) | 500 | 8037 | 17237 | 0.689 | 0.392 |
| % with Health Ins. Cov. | 92.7% | 12666 | 12608 | 0.629 | 0.396 |
| % with Internet Access | 81.8% | 12637 | 12637 | 0.974 | 0.429 |

**Table S6:** Summary of *post-matching* balance assessment and sample sizes across SDoH factors for treated and control groups.

| SDoH Factor | Max SMD | Mean SMD | Tr. Matched | Tr. Unmat. | Ct. Matched | Ct. Unmat. |
| --- | --- | --- | --- | --- | --- | --- |
| Median Household Income | 0.236 | 0.105 | 12555 | 21 | 3854 | 8720 |
| % Unemployed | 0.088 | 0.026 | 12544 | 31 | 5917 | 6658 |
| % Black or Afr. Am. Alone | 0.144 | 0.072 | 4932 | 0 | 1603 | 18615 |
| % Hispanic or Latino | 0.131 | 0.082 | 4018 | 0 | 2247 | 18885 |
| % with Bachelor or Higher | 0.113 | 0.069 | 12572 | 3 | 3981 | 8594 |
| Population Density (per mi<sup>2</sup>) | 0.181 | 0.114 | 8022 | 6 | 2836 | 14286 |
| % with Health Ins. Cov. | 0.067 | 0.030 | 8508 | 0 | 4339 | 12303 |
| % with Internet Access | 0.176 | 0.075 | 12575 | 0 | 3623 | 8952 |

**Table S7:** Balance assessment between unmatched high (Treated) and low (Control) ‘% Black or African American Alone’ Groups

| SDoH Factor | Means Treated | Means Control | Mean Diff. | Std. Mean Diff. |
| --- | --- | --- | --- | --- |
| distance | 0.590 | 0.100 | 0.490 | 1.606 |
| Median Household Income | 49810.138 | 63497.945 | -13687.808 | -0.680 |
| Unemployed | 0.045 | 0.031 | 0.015 | 0.628 |
| Hispanic or Latino | 0.125 | 0.095 | 0.030 | 0.201 |
| With Bachelor or Higher Degree Attained | 0.232 | 0.265 | -0.034 | -0.244 |
| Population Density (per sq mile) | 2975.905 | 1172.126 | 1803.778 | 0.245 |
| With Health Insurance Coverage | 0.883 | 0.918 | -0.034 | -0.575 |
| With Internet Access | 0.766 | 0.816 | -0.050 | -0.414 |
| Black or African American Alone | 0.337 | 0.021 | 0.316 | 1.517 |

**Table S8:** Balance assessment between matched high (Treated) and low (Control) ‘% Black or African American Alone’ groups

| SDoH Factor | Means Treated | Means Control | Mean Diff. | Std. Mean Diff. |
| --- | --- | --- | --- | --- |
| distance | 0.887 | 0.791 | 0.096 | 0.043 |
| Median Household Income | 49835.515 | 52509.535 | -2674.020 | -0.133 |
| Unemployed | 0.045 | 0.043 | 0.002 | 0.097 |
| Hispanic or Latino | 0.125 | 0.126 | -0.001 | -0.004 |
| With Bachelor or Higher Degree Attained | 0.232 | 0.240 | -0.008 | -0.059 |
| Population Density (per sq mile) | 2981.898 | 2250.314 | 731.584 | 0.099 |
| With Health Insurance Coverage | 0.884 | 0.881 | 0.003 | 0.044 |
| With Internet Access | 0.766 | 0.775 | -0.009 | -0.072 |
| Black or African American Alone | 0.337 | 0.041 | 0.297 | 1.425 |

**Table S9:** Balance assessment between unmatched high (Treated) and low (Control) ‘% Hispanic or Latino’ Groups

| SDoH Factor | Means Treated | Means Control | Mean Diff. | Std. Mean Diff. |
| --- | --- | --- | --- | --- |
| distance | 0.368 | 0.120 | 0.249 | 1.023 |
| Median Household Income | 57217.731 | 61502.769 | -4285.038 | -0.209 |
| Unemployed | 0.041 | 0.032 | 0.009 | 0.398 |
| Black or African American Alone | 0.099 | 0.080 | 0.019 | 0.145 |
| With Bachelor or Higher Degree Attained | 0.234 | 0.264 | -0.030 | -0.224 |
| Population Density (per sq mile) | 4021.757 | 1051.698 | 2970.060 | 0.333 |
| With Health Insurance Coverage | 0.864 | 0.920 | -0.056 | -0.751 |
| With Internet Access | 0.806 | 0.806 | -0.000 | -0.001 |
| Hispanic or Latino | 0.399 | 0.044 | 0.355 | 1.736 |

**Table S10:** Balance assessment between matched high (Treated) and low (Control) ‘% Hispanic or Latino’ groups

| SDoH Factor | Means Treated | Means Control | Mean Diff. | Std. Mean Diff. |
| --- | --- | --- | --- | --- |
| distance | -0.639 | -0.699 | 0.060 | 0.042 |
| Median Household Income | 57234.061 | 58095.893 | -861.832 | -0.042 |
| Unemployed | 0.041 | 0.038 | 0.002 | 0.105 |
| Black or African American Alone | 0.099 | 0.085 | 0.014 | 0.111 |
| With Bachelor or Higher Degree Attained | 0.234 | 0.239 | -0.006 | -0.042 |
| Population Density (per sq mile) | 4029.507 | 2986.171 | 1043.336 | 0.117 |
| With Health Insurance Coverage | 0.864 | 0.865 | -0.001 | -0.018 |
| With Internet Access | 0.806 | 0.817 | -0.010 | -0.096 |
| Hispanic or Latino | 0.399 | 0.086 | 0.313 | 1.530 |

**Table S11:**
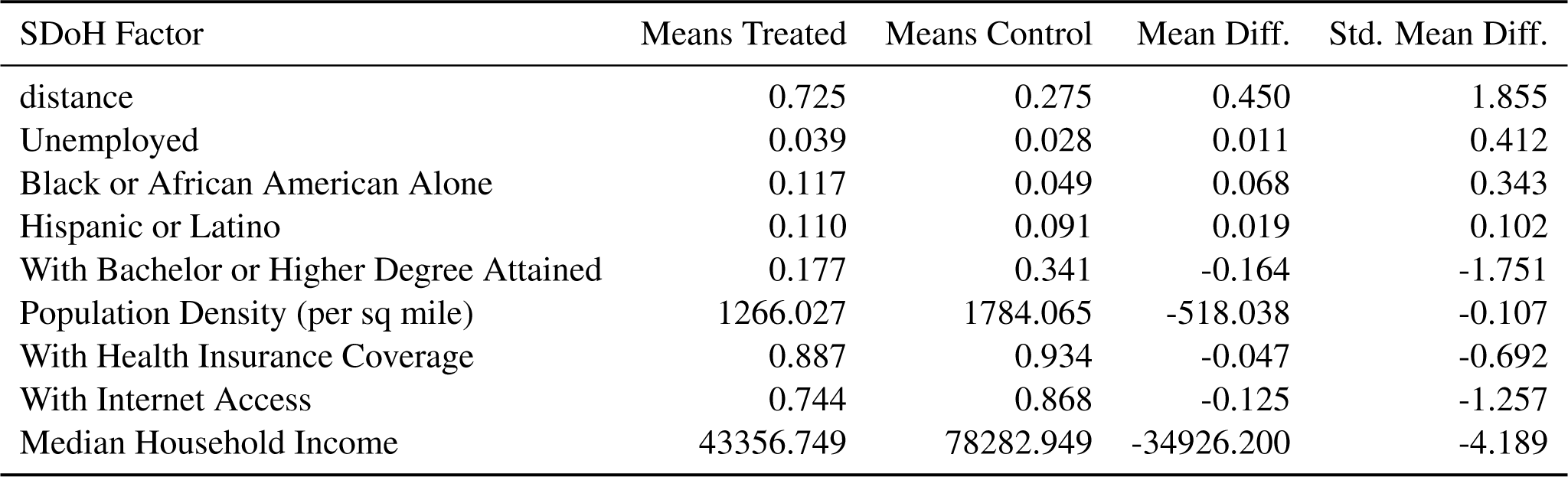
Balance assessment between unmatched high (Control) and low (Treated) ‘Median Household Income’ Groups

**Table S12:** Balance assessment between matched high (Control) and low (Treated) ‘Median Household Income’ groups

| SDoH Factor | Means Treated | Means Control | Mean Diff. | Std. Mean Diff. |
| --- | --- | --- | --- | --- |
| distance | 1.567 | 1.449 | 0.119 | 0.060 |
| Unemployed | 0.039 | 0.036 | 0.003 | 0.113 |
| Black or African American Alone | 0.117 | 0.092 | 0.025 | 0.126 |
| Hispanic or Latino | 0.111 | 0.091 | 0.020 | 0.107 |
| With Bachelor or Higher Degree Attained | 0.177 | 0.180 | -0.003 | -0.029 |
| Population Density (per sq mile) | 1272.878 | 747.235 | 525.643 | 0.108 |
| With Health Insurance Coverage | 0.888 | 0.889 | -0.001 | -0.021 |
| With Internet Access | 0.745 | 0.746 | -0.001 | -0.011 |
| Median Household Income | 43401.084 | 62615.121 | -19214.037 | -2.306 |

**Table S13:**
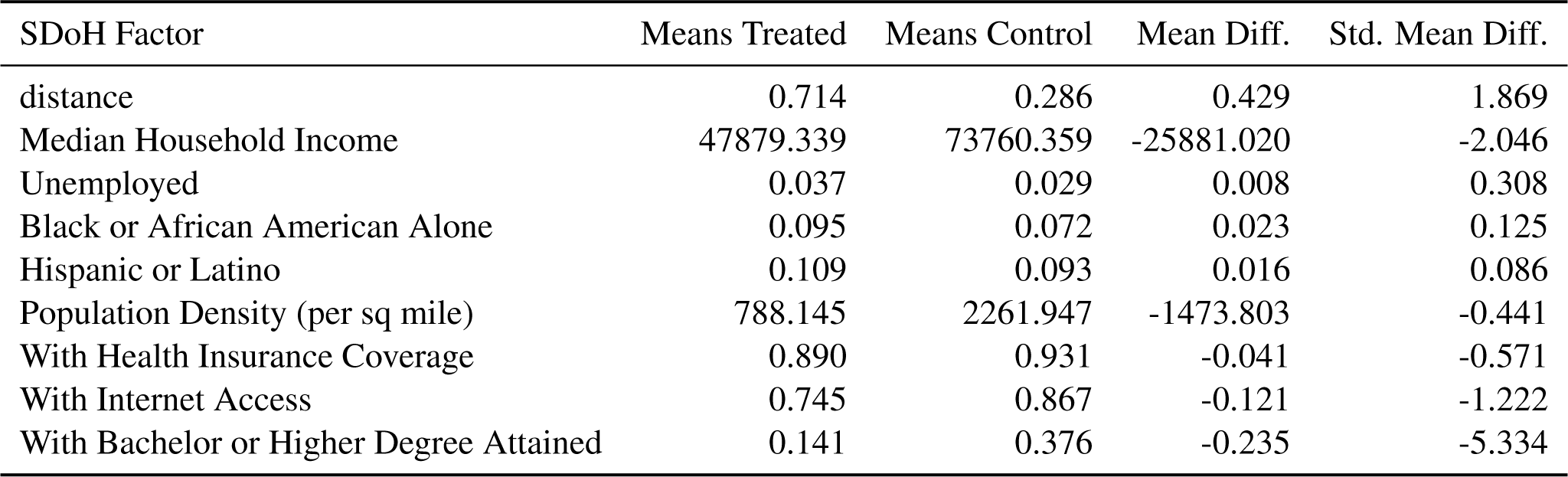
Balance assessment between unmatched high (Control) and low (Treated) ‘% with Bachelor or Higher Degree Attained’ Groups

**Table S14:** Balance assessment between matched high (Control) and low (Treated) ‘% with Bachelor or Higher Degree Attained’ groups

| SDoH Factor | Means Treated | Means Control | Mean Diff. | Std. Mean Diff. |
| --- | --- | --- | --- | --- |
| distance | 1.336 | 1.242 | 0.094 | 0.058 |
| Median Household Income | 47895.952 | 48454.763 | -558.811 | -0.044 |
| Unemployed | 0.037 | 0.036 | 0.002 | 0.073 |
| Black or African American Alone | 0.095 | 0.084 | 0.011 | 0.060 |
| Hispanic or Latino | 0.109 | 0.097 | 0.013 | 0.066 |
| Population Density (per sq mile) | 793.735 | 751.143 | 42.592 | 0.013 |
| With Health Insurance Coverage | 0.891 | 0.897 | -0.007 | -0.097 |
| With Internet Access | 0.746 | 0.751 | -0.005 | -0.050 |
| With Bachelor or Higher Degree Attained | 0.142 | 0.269 | -0.128 | -2.903 |

**Table S15:**
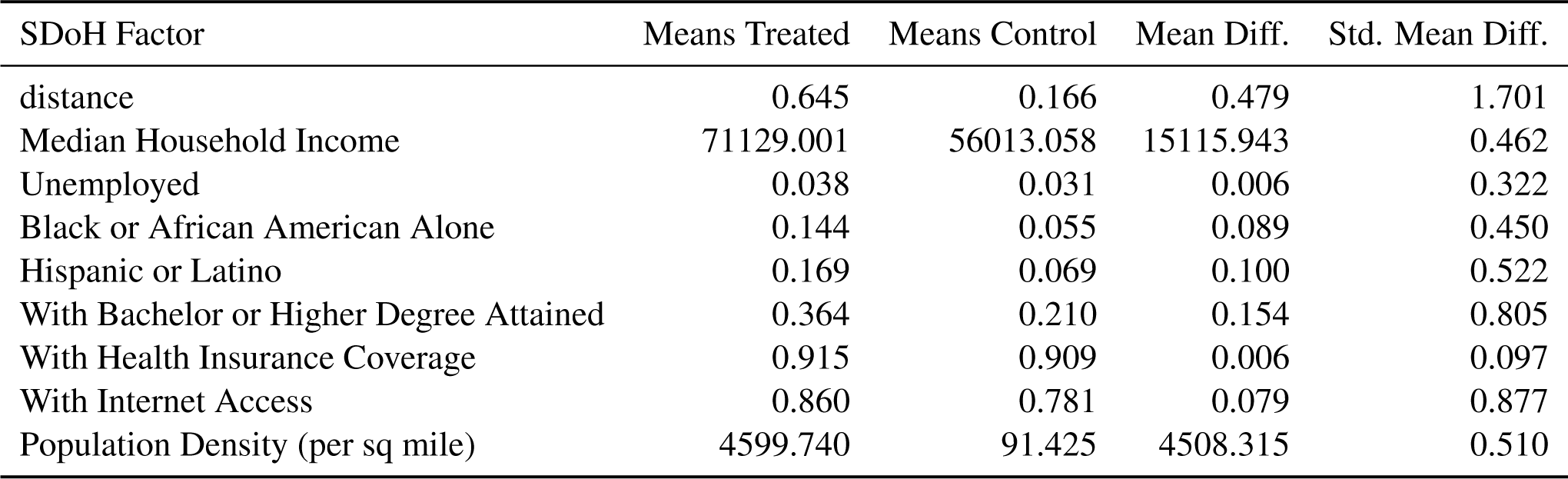
Balance assessment between unmatched high (Treated) and low (Control) ‘Population Density (per sq mile)’ Groups

**Table S16:**
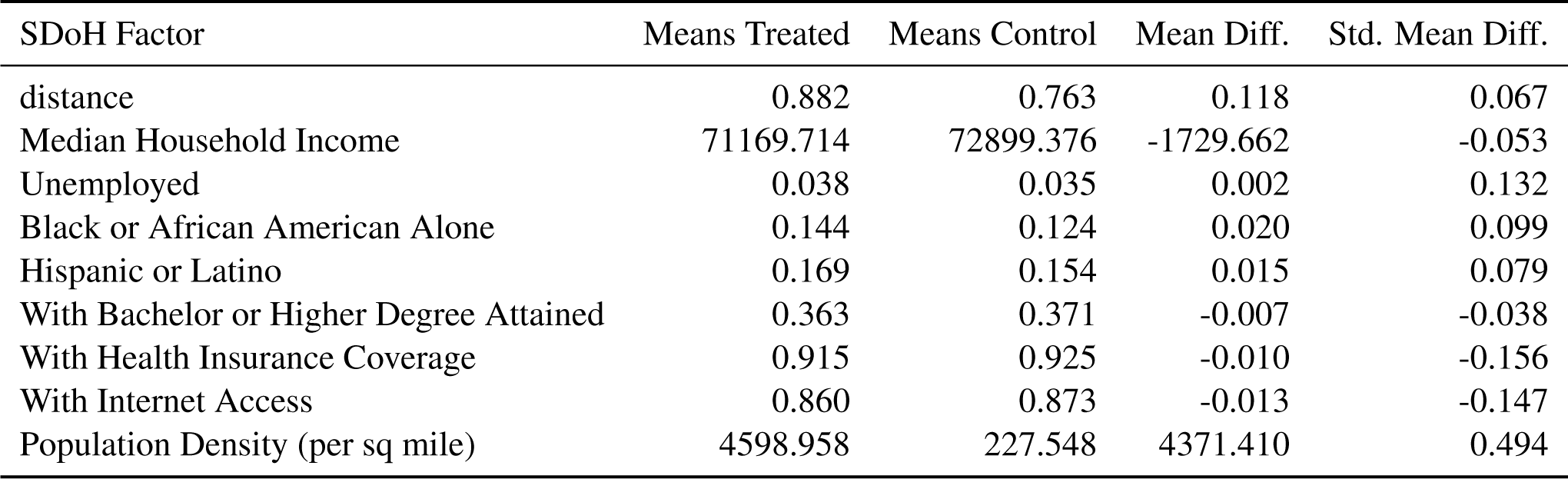
Balance assessment between matched high (Treated) and low (Control) ‘Population Density (per sq mile)’ groups

**Table S17:** Balance assessment between unmatched high (Treated) and low (Control) ‘% Unemployed’ Groups

| SDoH Factor | Means Treated | Means Control | Mean Diff. | Std. Mean Diff. |
| --- | --- | --- | --- | --- |
| distance | 0.581 | 0.419 | 0.162 | 0.781 |
| Median Household Income | 55178.441 | 66461.257 | -11282.816 | -0.503 |
| Black or African American Alone | 0.125 | 0.042 | 0.083 | 0.421 |
| Hispanic or Latino | 0.132 | 0.070 | 0.062 | 0.328 |
| With Bachelor or Higher Degree Attained | 0.230 | 0.288 | -0.058 | -0.416 |
| Population Density (per sq mile) | 2062.020 | 988.072 | 1073.948 | 0.173 |
| With Health Insurance Coverage | 0.898 | 0.924 | -0.026 | -0.404 |
| With Internet Access | 0.791 | 0.821 | -0.029 | -0.272 |
| Unemployed | 0.049 | 0.018 | 0.030 | 1.401 |

**Table S18:** Balance assessment between matched high (Treated) and low (Control) ‘% Unemployed’ groups

| SDoH Factor | Means Treated | Means Control | Mean Diff. | Std. Mean Diff. |
| --- | --- | --- | --- | --- |
| distance | 0.460 | 0.423 | 0.037 | 0.032 |
| Median Household Income | 55192.003 | 55360.831 | -168.828 | -0.008 |
| Black or African American Alone | 0.124 | 0.122 | 0.002 | 0.010 |
| Hispanic or Latino | 0.131 | 0.125 | 0.006 | 0.030 |
| With Bachelor or Higher Degree Attained | 0.230 | 0.225 | 0.004 | 0.030 |
| Population Density (per sq mile) | 1902.049 | 1476.827 | 425.222 | 0.069 |
| With Health Insurance Coverage | 0.898 | 0.898 | -0.001 | -0.010 |
| With Internet Access | 0.791 | 0.791 | 0.001 | 0.006 |
| Unemployed | 0.048 | 0.021 | 0.028 | 1.303 |

**Table S19:** Balance assessment between unmatched high (Control) and low (Treated) ‘% with Health Insurance Coverage’ Groups

| SDoH Factor | Means Treated | Means Control | Mean Diff. | Std. Mean Diff. |
| --- | --- | --- | --- | --- |
| distance | 0.528 | 0.242 | 0.286 | 1.140 |
| Median Household Income | 48183.387 | 67291.556 | -19108.168 | -1.336 |
| Unemployed | 0.040 | 0.030 | 0.010 | 0.374 |
| Black or African American Alone | 0.133 | 0.058 | 0.075 | 0.378 |
| Hispanic or Latino | 0.167 | 0.067 | 0.101 | 0.458 |
| With Bachelor or Higher Degree Attained | 0.187 | 0.296 | -0.109 | -1.076 |
| Population Density (per sq mile) | 1529.525 | 1522.752 | 6.774 | 0.001 |
| With Internet Access | 0.752 | 0.834 | -0.082 | -0.737 |
| With Health Insurance Coverage | 0.841 | 0.947 | -0.106 | -1.770 |

**Table S20:**
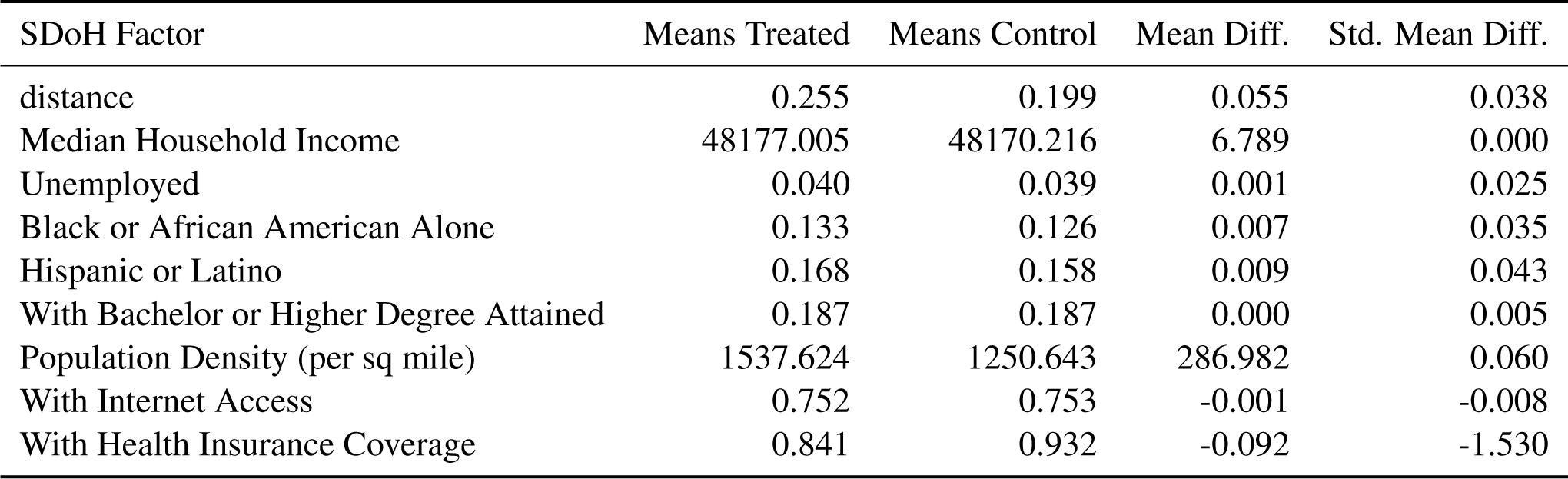
Balance assessment between matched high (Control) and low (Treated) ‘% with Health Insurance Coverage’ groups

| SDoH Factor | Means Treated | Means Control | Mean Diff. | Std. Mean Diff. |
| --- | --- | --- | --- | --- |
| distance | 0.255 | 0.199 | 0.055 | 0.038 |
| Median Household Income | 48177.005 | 48170.216 | 6.789 | 0.000 |
| Unemployed | 0.040 | 0.039 | 0.001 | 0.025 |
| Black or African American Alone | 0.133 | 0.126 | 0.007 | 0.035 |
| Hispanic or Latino | 0.168 | 0.158 | 0.009 | 0.043 |
| With Bachelor or Higher Degree Attained | 0.187 | 0.187 | 0.000 | 0.005 |
| Population Density (per sq mile) | 1537.624 | 1250.643 | 286.982 | 0.060 |
| With Internet Access | 0.752 | 0.753 | -0.001 | -0.008 |
| With Health Insurance Coverage | 0.841 | 0.932 | -0.092 | -1.530 |

**Table S21:** Balance assessment between unmatched high (Control) and low (Treated) ‘% with Internet Access’ Groups

| SDoH Factor | Means Treated | Means Control | Mean Diff. | Std. Mean Diff. |
| --- | --- | --- | --- | --- |
| distance | 0.749 | 0.251 | 0.497 | 2.260 |
| Median Household Income | 46716.098 | 74923.599 | -28207.501 | -2.274 |
| Unemployed | 0.037 | 0.030 | 0.007 | 0.256 |
| Black or African American Alone | 0.107 | 0.060 | 0.047 | 0.244 |
| Hispanic or Latino | 0.102 | 0.099 | 0.003 | 0.019 |
| With Bachelor or Higher Degree Attained | 0.168 | 0.350 | -0.182 | -2.326 |
| Population Density (per sq mile) | 1003.589 | 2046.502 | -1042.913 | -0.225 |
| With Health Insurance Coverage | 0.889 | 0.932 | -0.043 | -0.603 |
| With Internet Access | 0.723 | 0.889 | -0.166 | -1.980 |

**Table S22:** Balance assessment between matched high (Control) and low (Treated) ‘% with Internet Access’ groups

| SDoH Factor | Means Treated | Means Control | Mean Diff. | Std. Mean Diff. |
| --- | --- | --- | --- | --- |
| distance | 1.436 | 1.234 | 0.202 | 0.136 |
| Median Household Income | 46721.584 | 48710.356 | -1988.773 | -0.160 |
| Unemployed | 0.037 | 0.035 | 0.001 | 0.056 |
| Black or African American Alone | 0.107 | 0.096 | 0.011 | 0.059 |
| Hispanic or Latino | 0.103 | 0.093 | 0.009 | 0.050 |
| With Bachelor or Higher Degree Attained | 0.168 | 0.173 | -0.005 | -0.066 |
| Population Density (per sq mile) | 1009.178 | 898.001 | 111.178 | 0.024 |
| With Health Insurance Coverage | 0.889 | 0.893 | -0.003 | -0.049 |
| With Internet Access | 0.724 | 0.854 | -0.130 | -1.559 |

**Figure S1:**
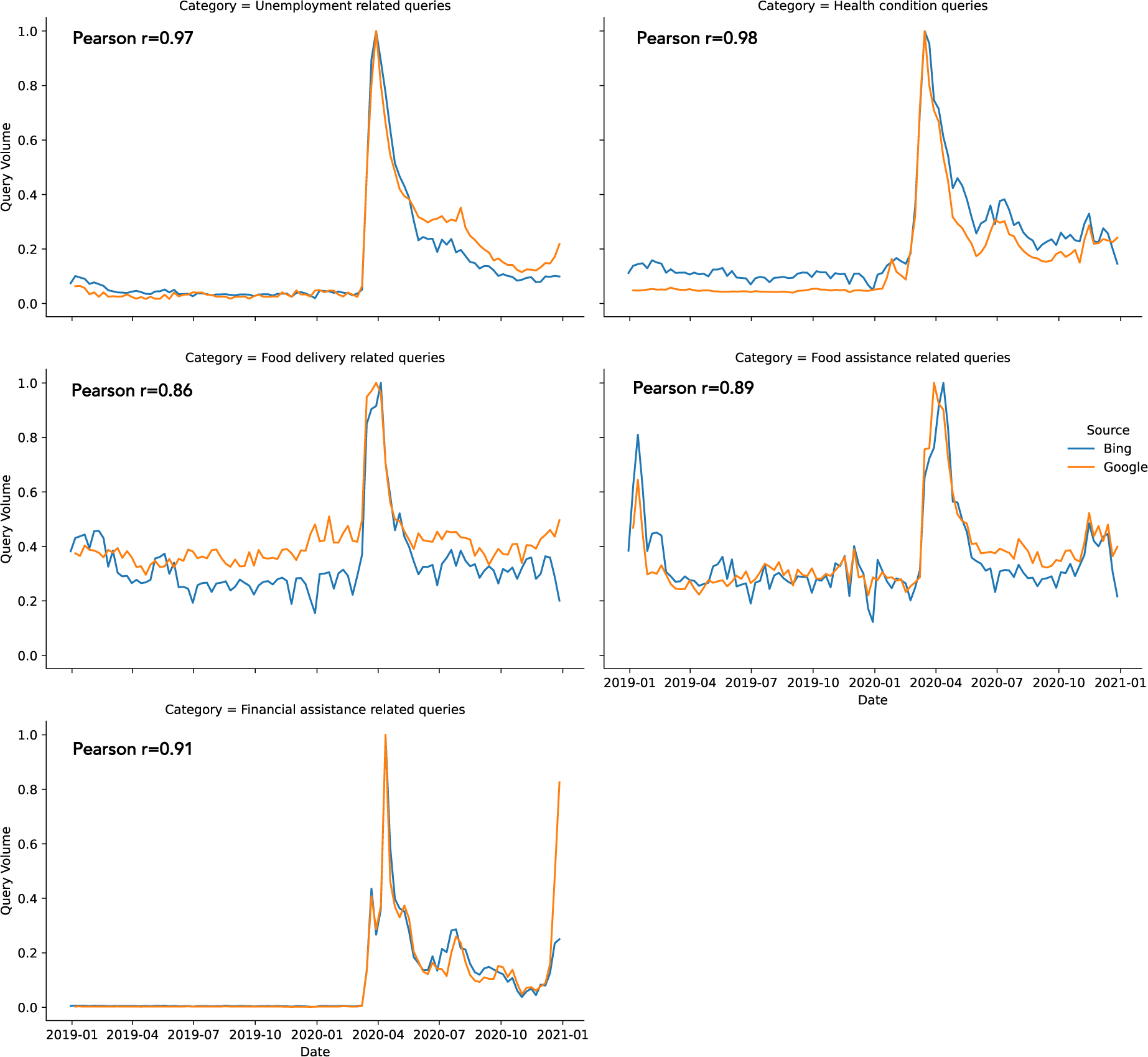
Normalized query volumes across five categories in years 2019 and 2020 across Bing and Google.

**Figure S2:**
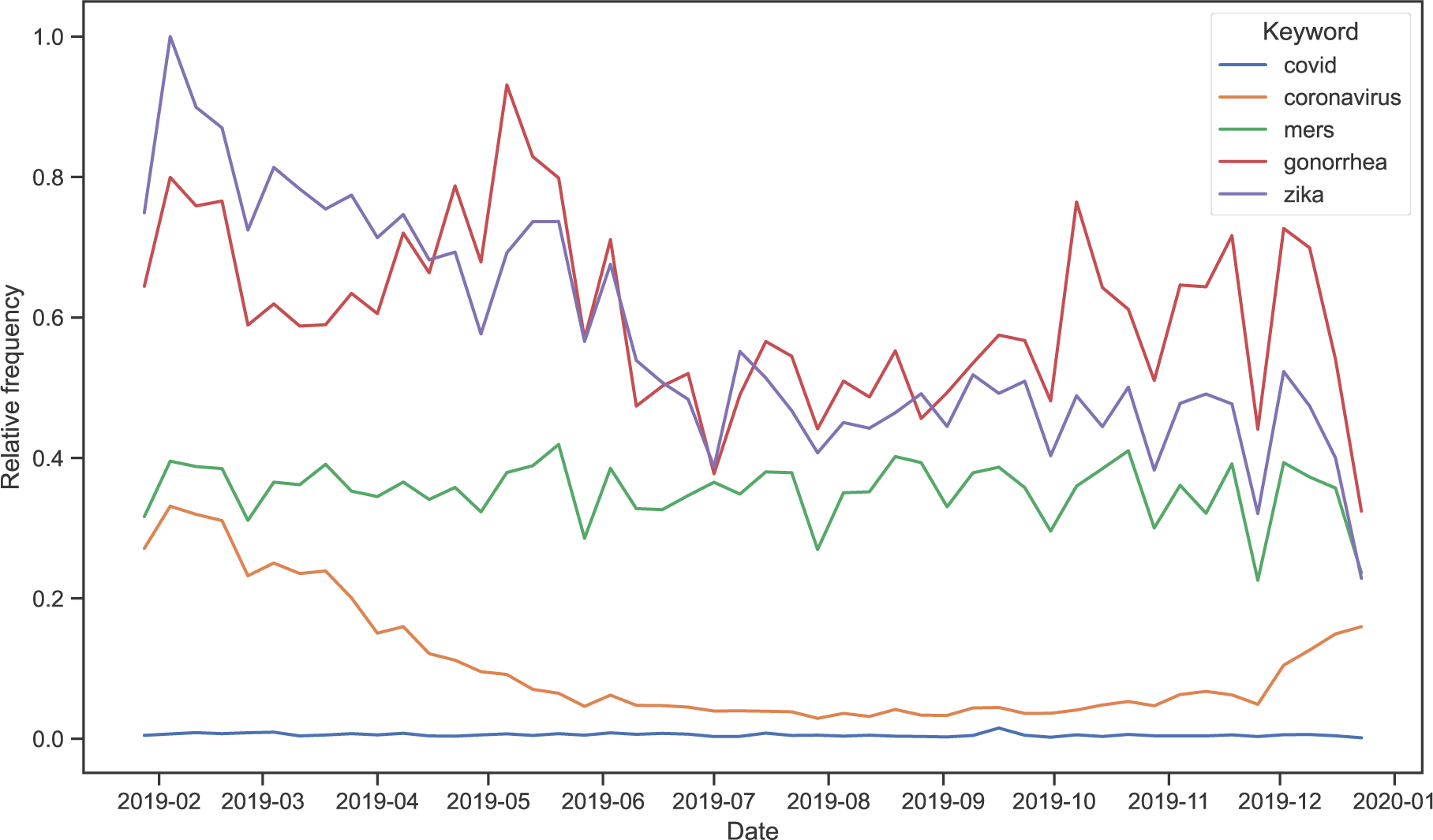
Frequency of search keywords “covid”, “coronavirus”, “mers”, “gonorrhea”, and “zika” in 2019, scaled to the maximum frequency within the data presented in the figure.

**Figure S3:**
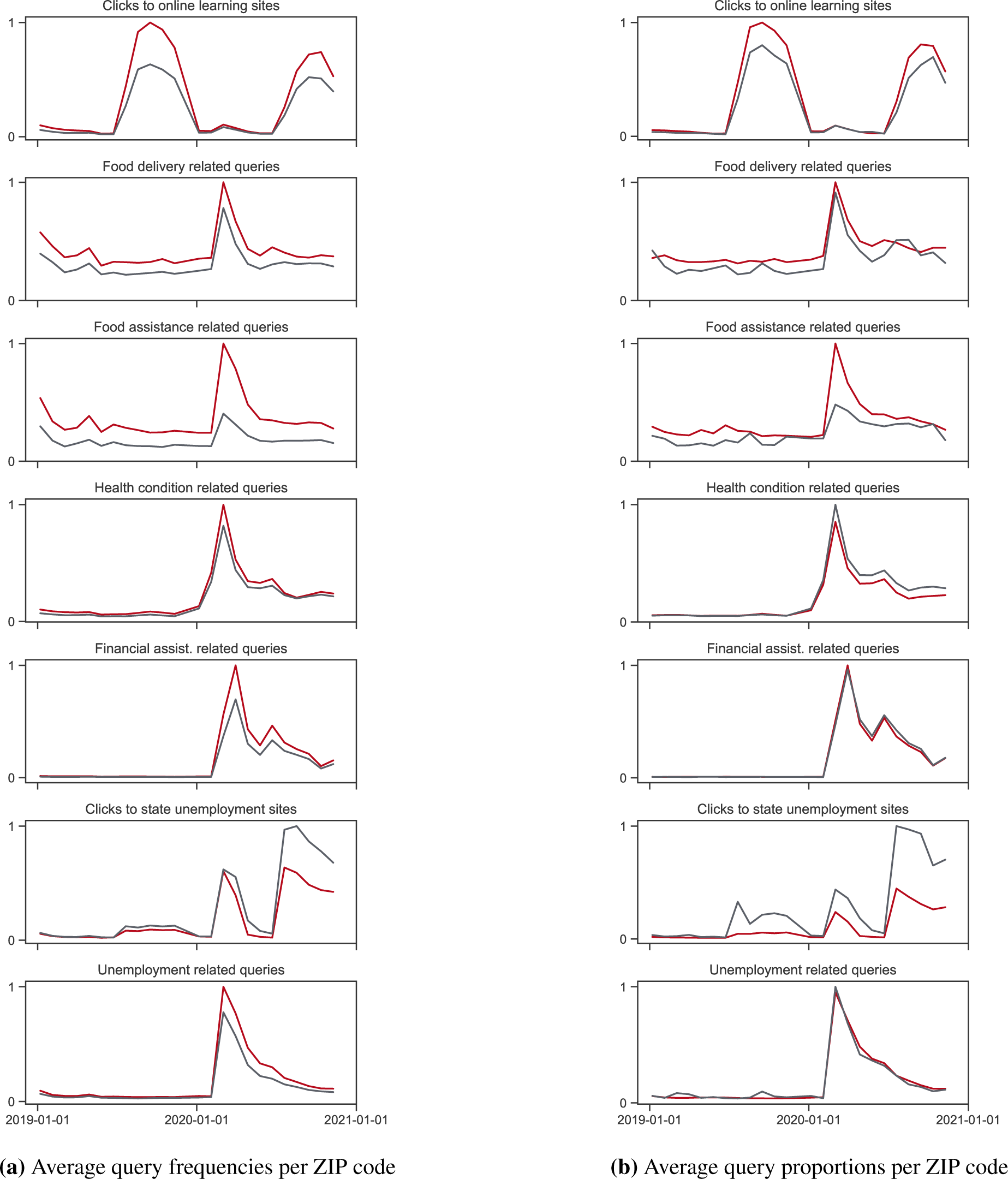
(a) Average query frequencies per ZIP code across seven search categories, and (b) average proportion of total queries per ZIP code across seven search categories. Query frequencies and proportions are scaled to the maximum of 1 within each figure. Red lines indicate ZIP codes with % of the population with Black or African American alone *≥* 12%, and gray lines indicate ZIP codes with % of the population with Black or African American alone < 12%.

**Figure S4:**
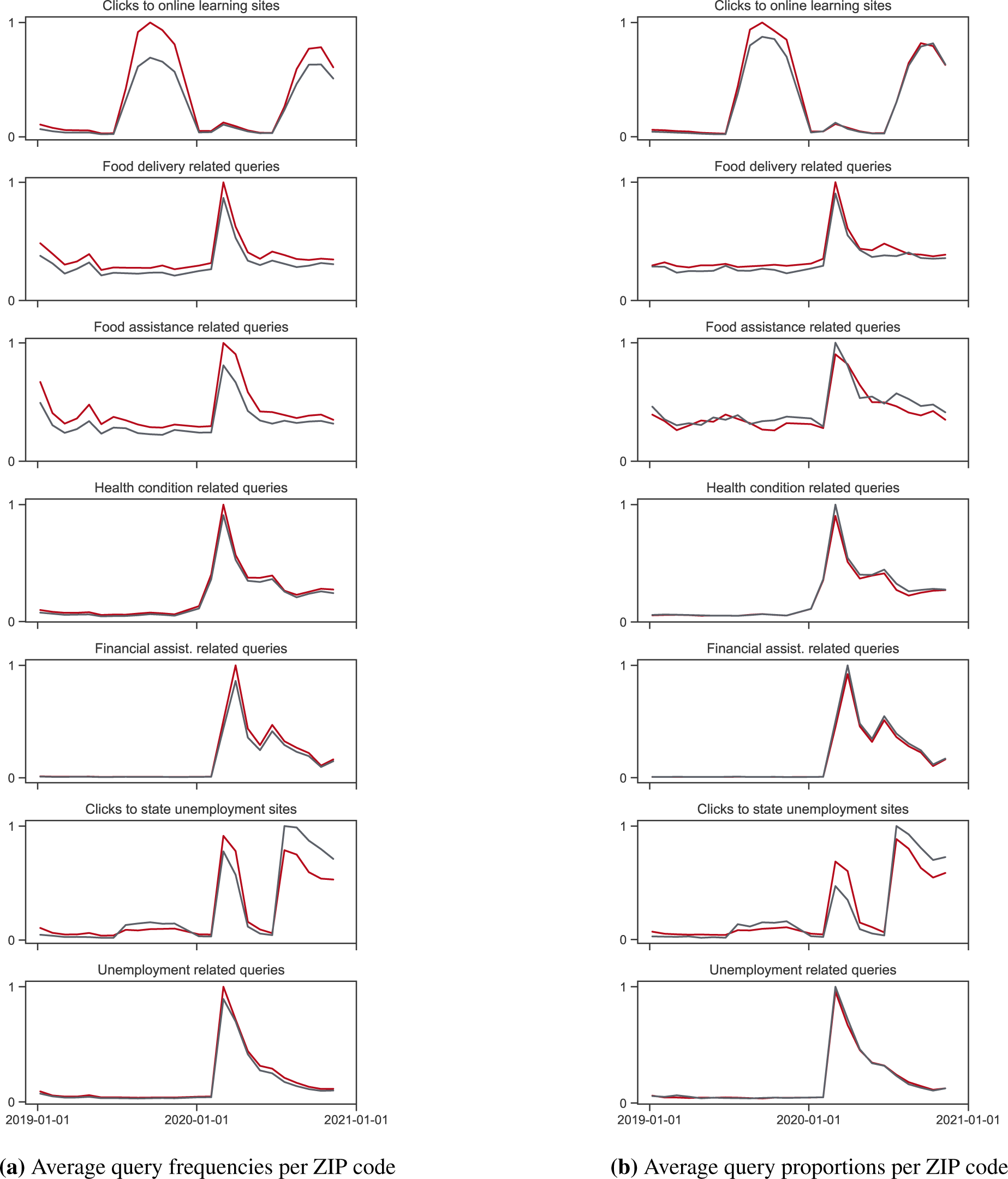
(a) Average query frequencies per ZIP code across seven search categories, and (b) average proportion of total queries per ZIP code across seven search categories. Query frequencies and proportions are scaled to the maximum of 1 within each figure. Red lines indicate ZIP codes with % of the population with Hispanic origin *≥* 18%, and gray lines indicate ZIP codes with % of the population with Hispanic origin < 18%.

**Figure S5:**
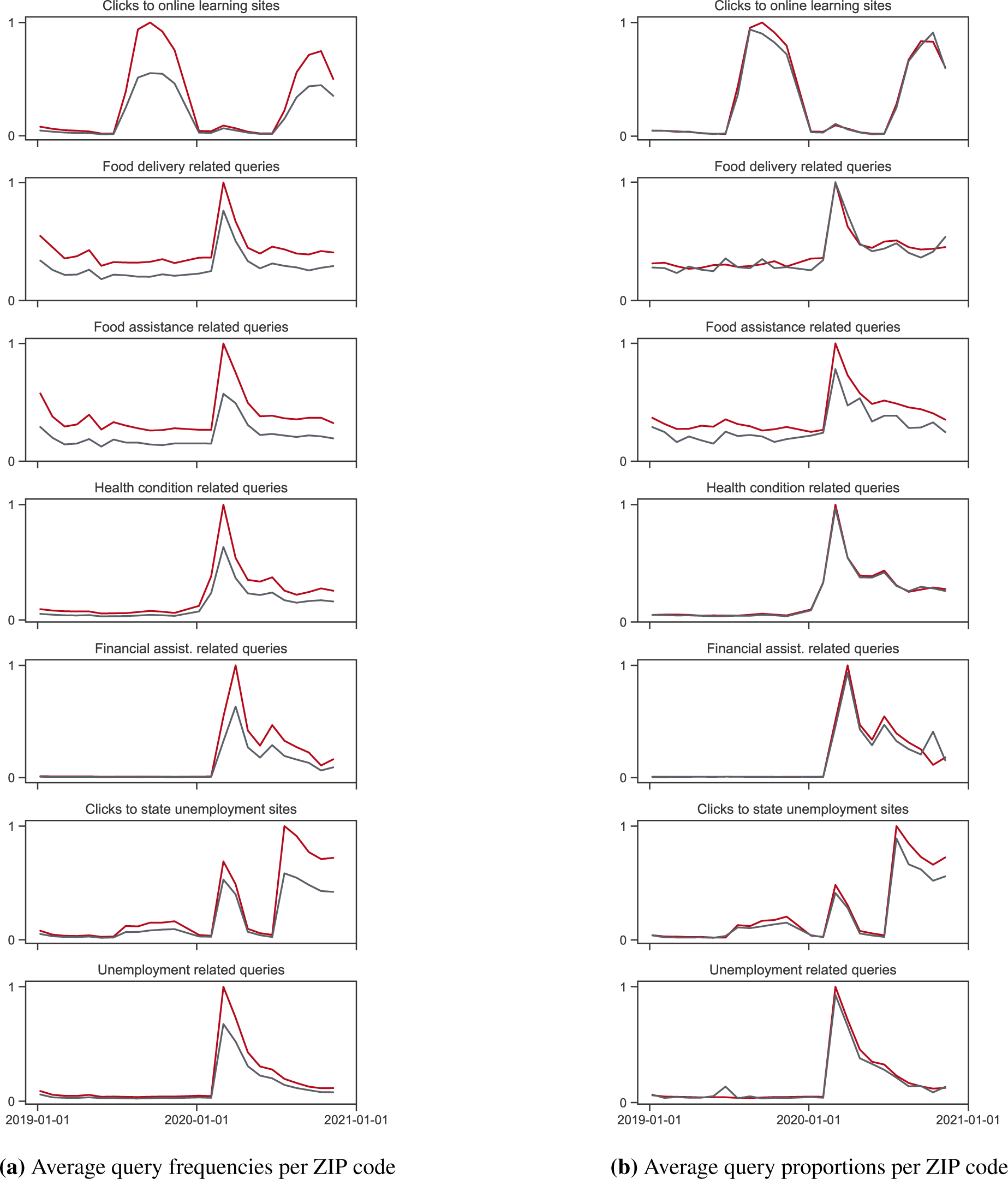
(a) Average query frequencies per ZIP code across seven search categories, and (b) average proportion of total queries per ZIP code across seven search categories. Query frequencies and proportions are scaled to the maximum of 1 within each figure. Red lines indicate ZIP codes with median household income *≤* $55,224, and gray lines indicate ZIP codes with median household income > $55,224.

**Figure S6:**
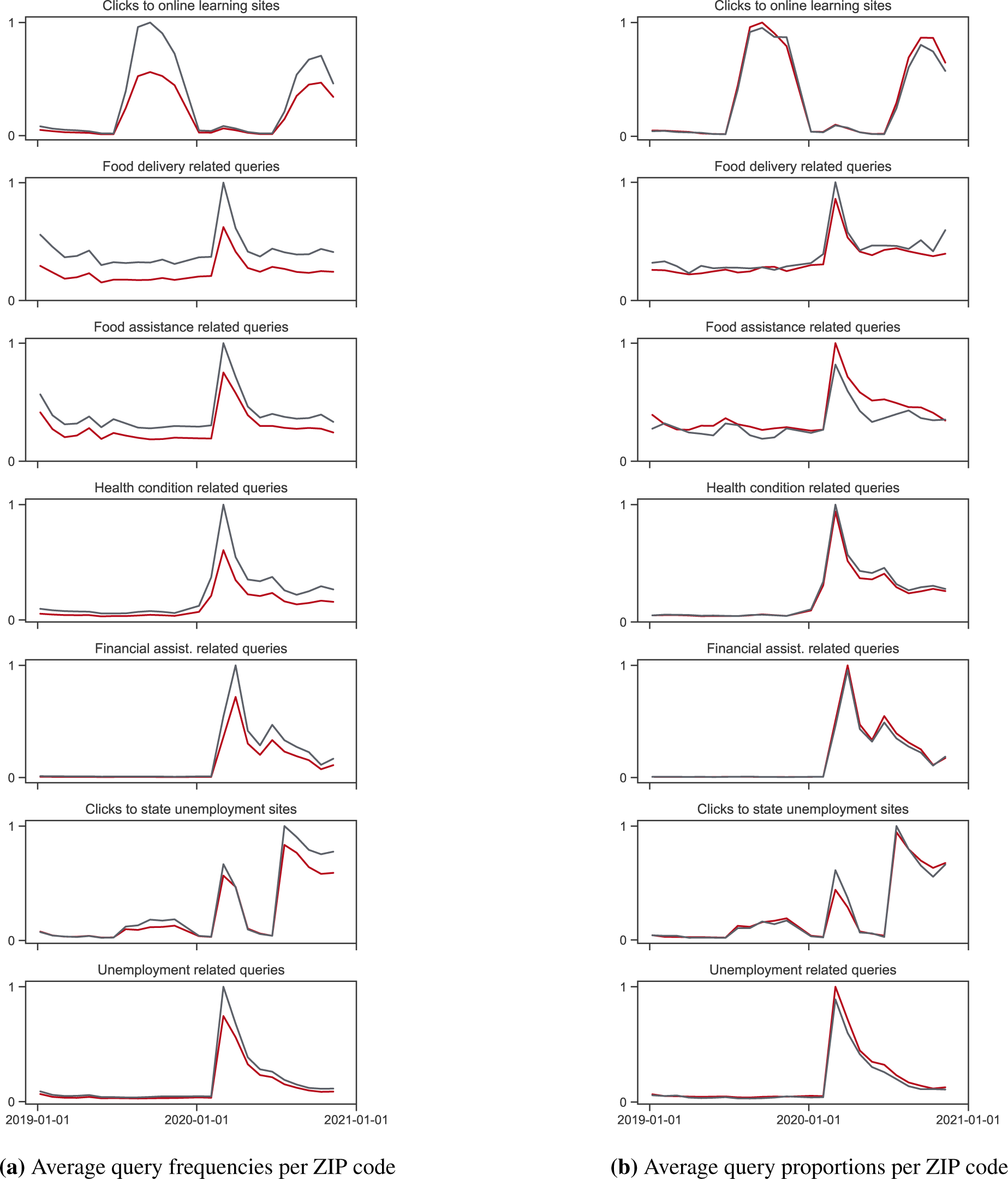
(a) Average query frequencies per ZIP code across seven search categories, and (b) average proportion of total queries per ZIP code across seven search categories. Query frequencies and proportions are scaled to the maximum of 1 within each figure. Red lines indicate ZIP codes with % of the population with Bachelor’s degree or higher degrees *≤* 21%, and gray lines indicate ZIP codes with Bachelor’s degree or higher degrees > 21%.

**Figure S7:**
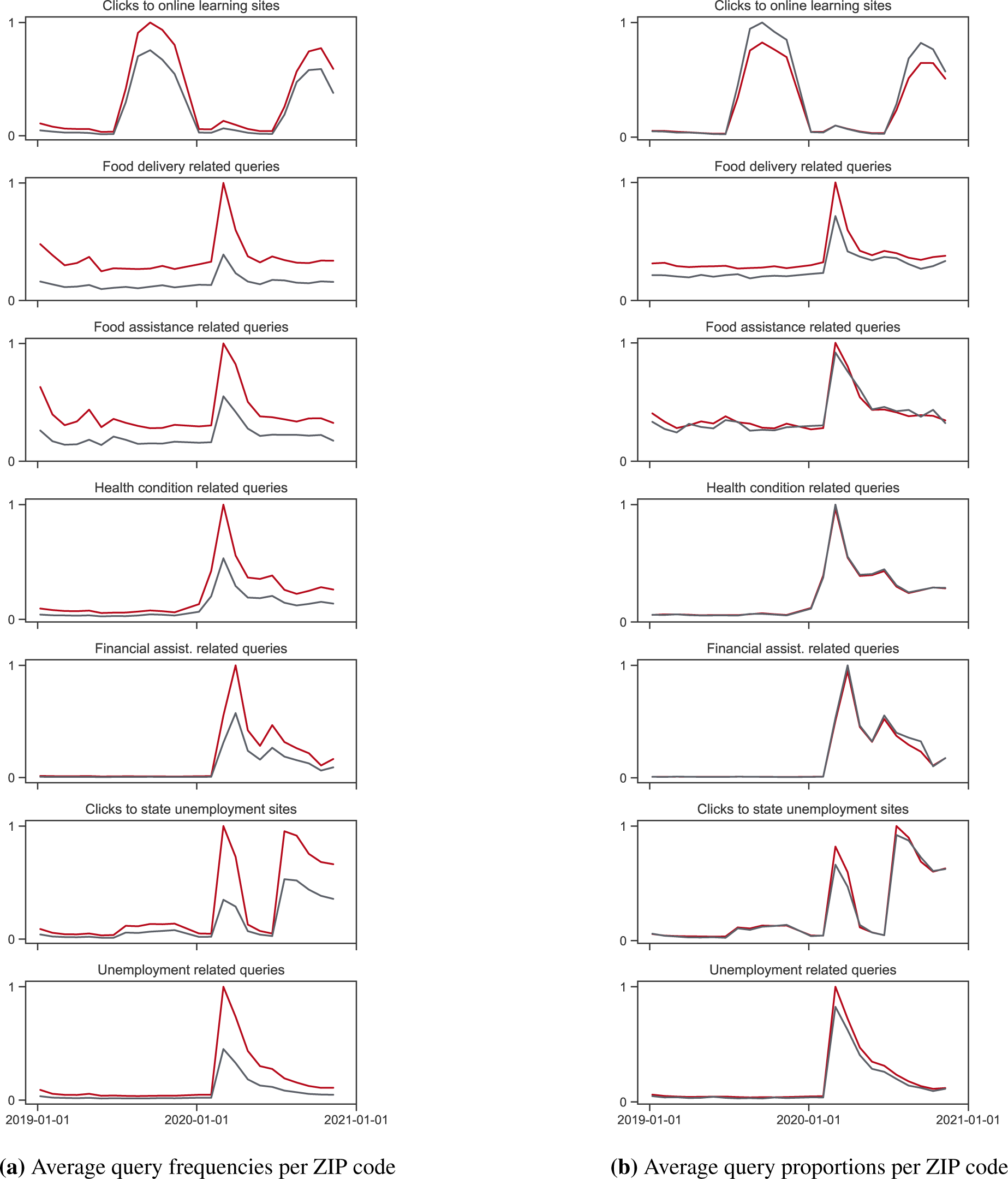
(a) Average query frequencies per ZIP code across seven search categories, and (b) average proportion of total queries per ZIP code across seven search categories. Query frequencies and proportions are scaled to the maximum of 1 within each figure. Red lines indicate ZIP codes with population density *≥* 500 people per square mile, and gray lines indicate ZIP codes with internet access < 500 people per square mile.

**Figure S8:**
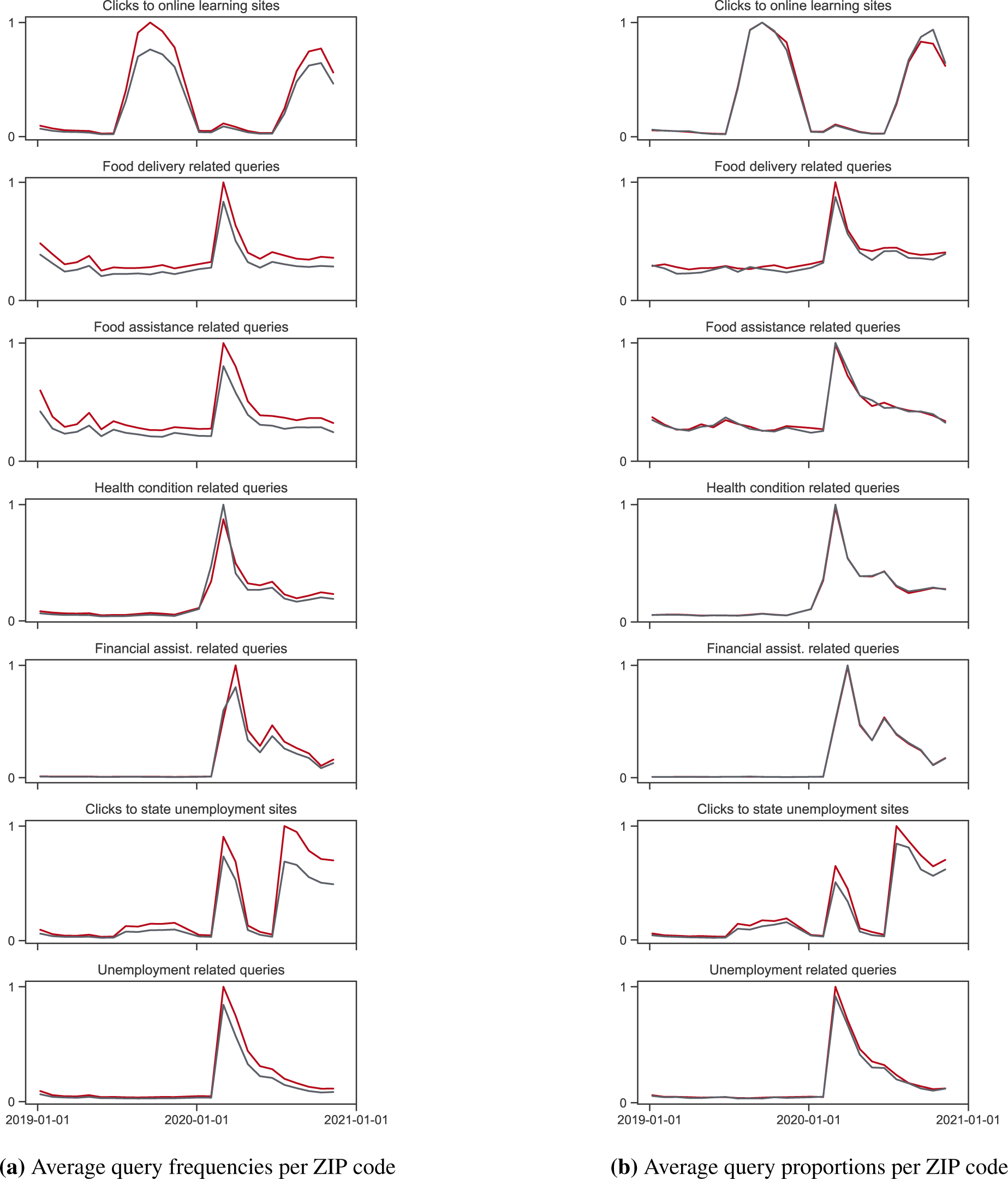
(a) Average query frequencies per ZIP code across seven search categories, and (b) average proportion of total queries per ZIP code across seven search categories. Query frequencies and proportions are scaled to the maximum of 1 within each figure. Red lines indicate ZIP codes with % of the population unemployed *≥* 3%, and gray lines indicate ZIP codes with % of the population unemployed < 3%.

**Figure S9:**
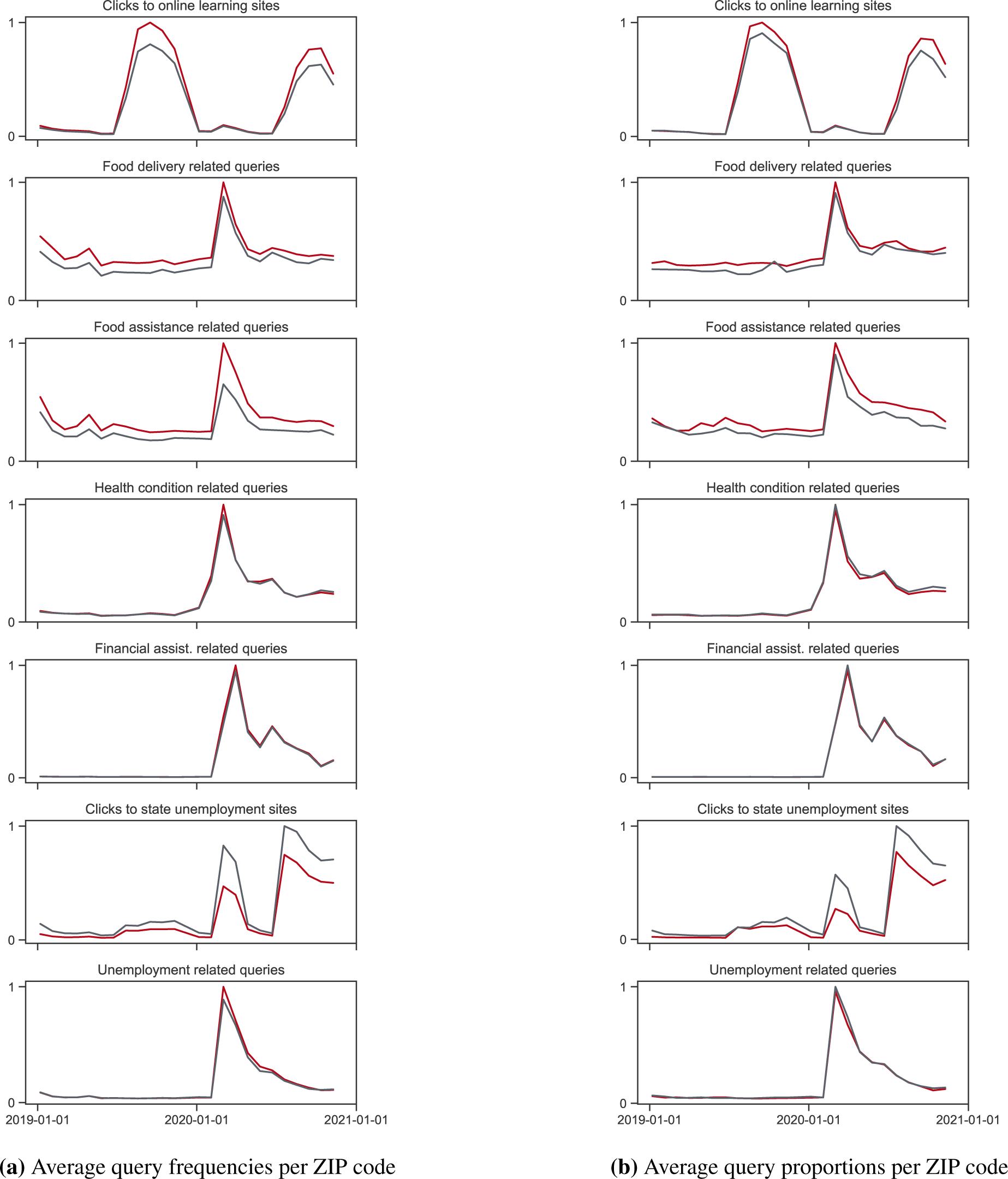
(a) Average query frequencies per ZIP code across seven search categories, and (b) average proportion of total queries per ZIP code across seven search categories. Query frequencies and proportions are scaled to the maximum of 1 within each figure. Red lines indicate ZIP codes with % of the population with health insurance coverage *≤* 93%, and gray lines indicate ZIP codes with health insurance coverage > 93%.

**Figure S10:**
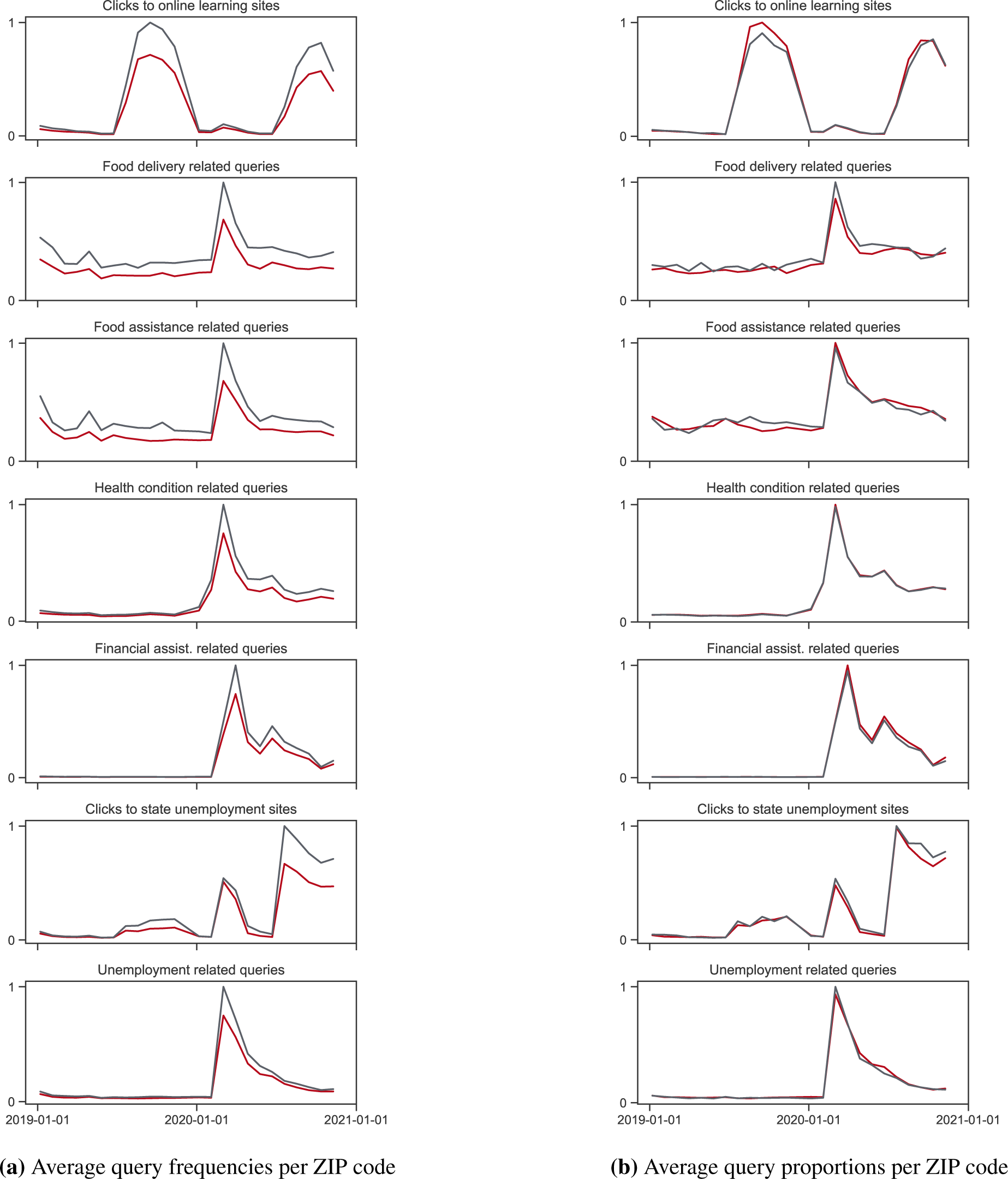
(a) Average query frequencies per ZIP code across seven search categories, and (b) average proportion of total queries per ZIP code across seven search categories. Query frequencies and proportions are scaled to the maximum of 1 within each figure. Red lines indicate ZIP codes with % of the population with internet access *≤* 82%, and gray lines indicate ZIP codes with internet access > 82%.

**Figure S11:**
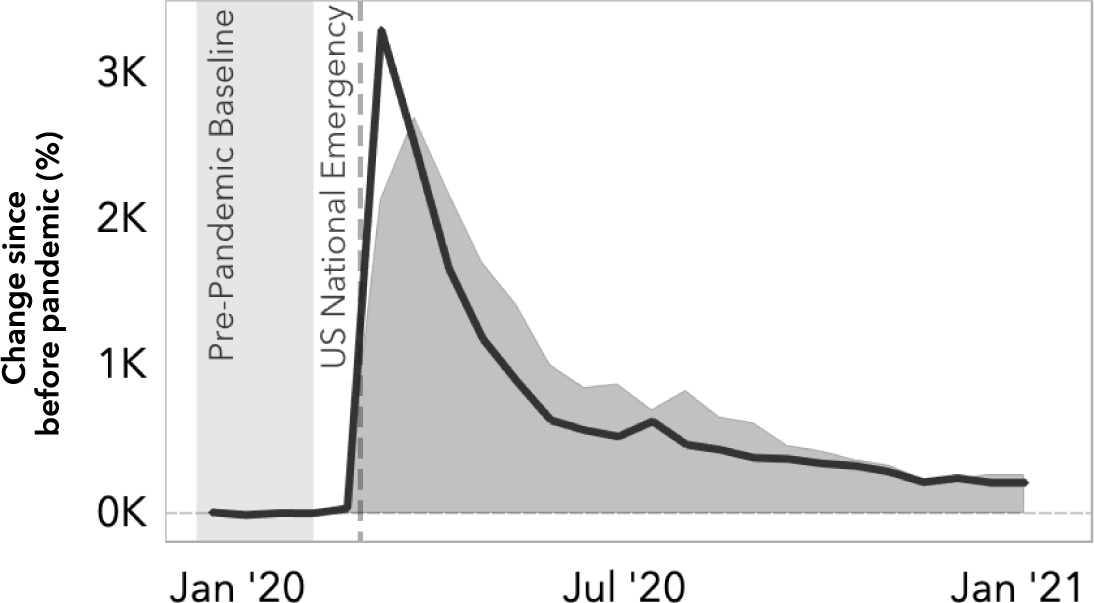
Percent change in the unemployment related queries in Bing (shaded) and the reported unemployment claims from the US Department of Labor (line, https://oui.doleta.gov/unemploy/claims. asp) compared to pre-pandemic baseline.

**Figure S12:**
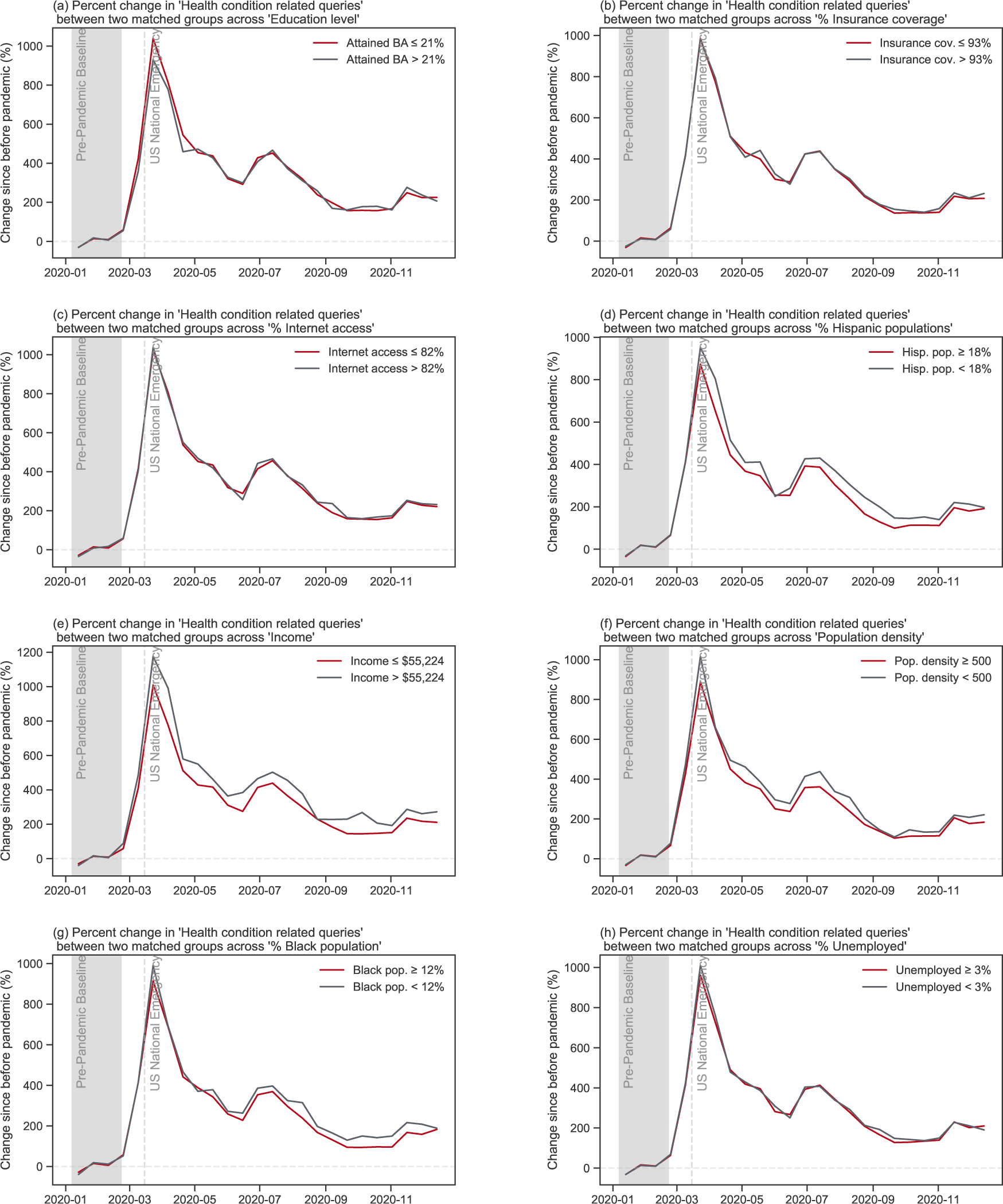
Percent change in ‘Health condition related queries’ between two matched groups across eight SDoH factors.

**Figure S13:**
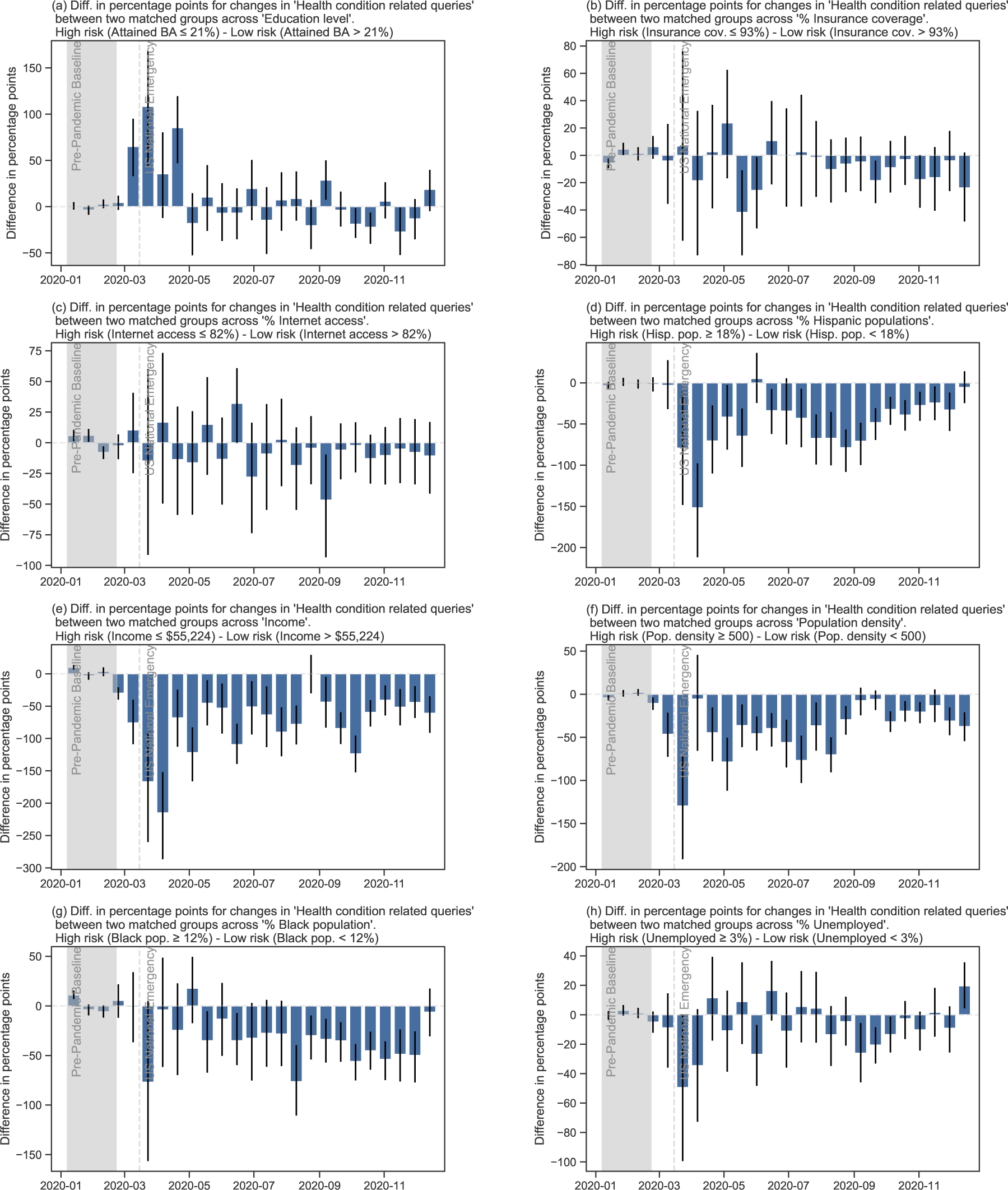
Differences in percentage points for changes in ‘Health condition related queries’ between two matched groups across eight SDoH factors.

**Figure S14:**
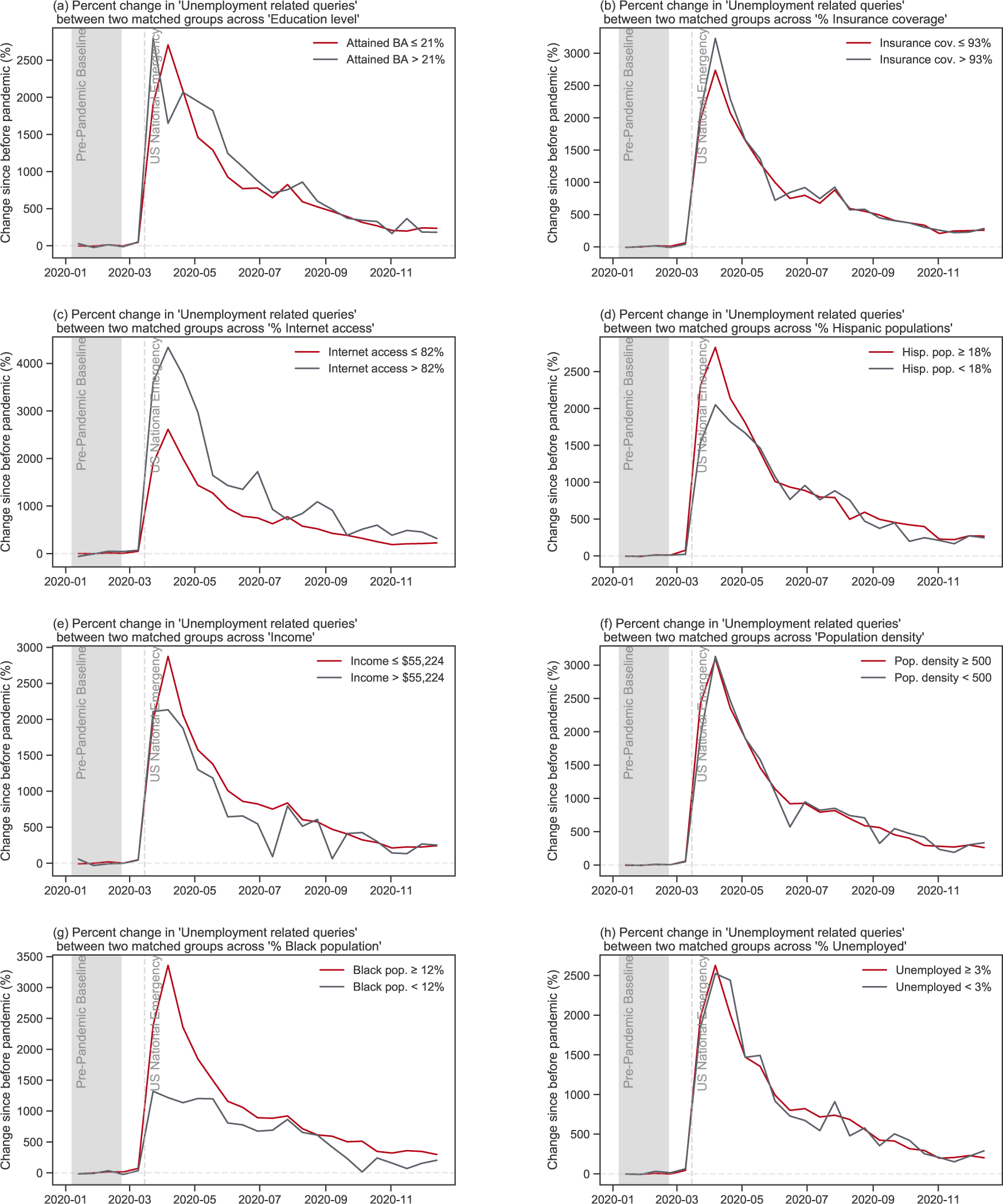
Percent change in ‘Unemployment related queries’ between two matched groups across eight SDoH factors.

**Figure S15:**
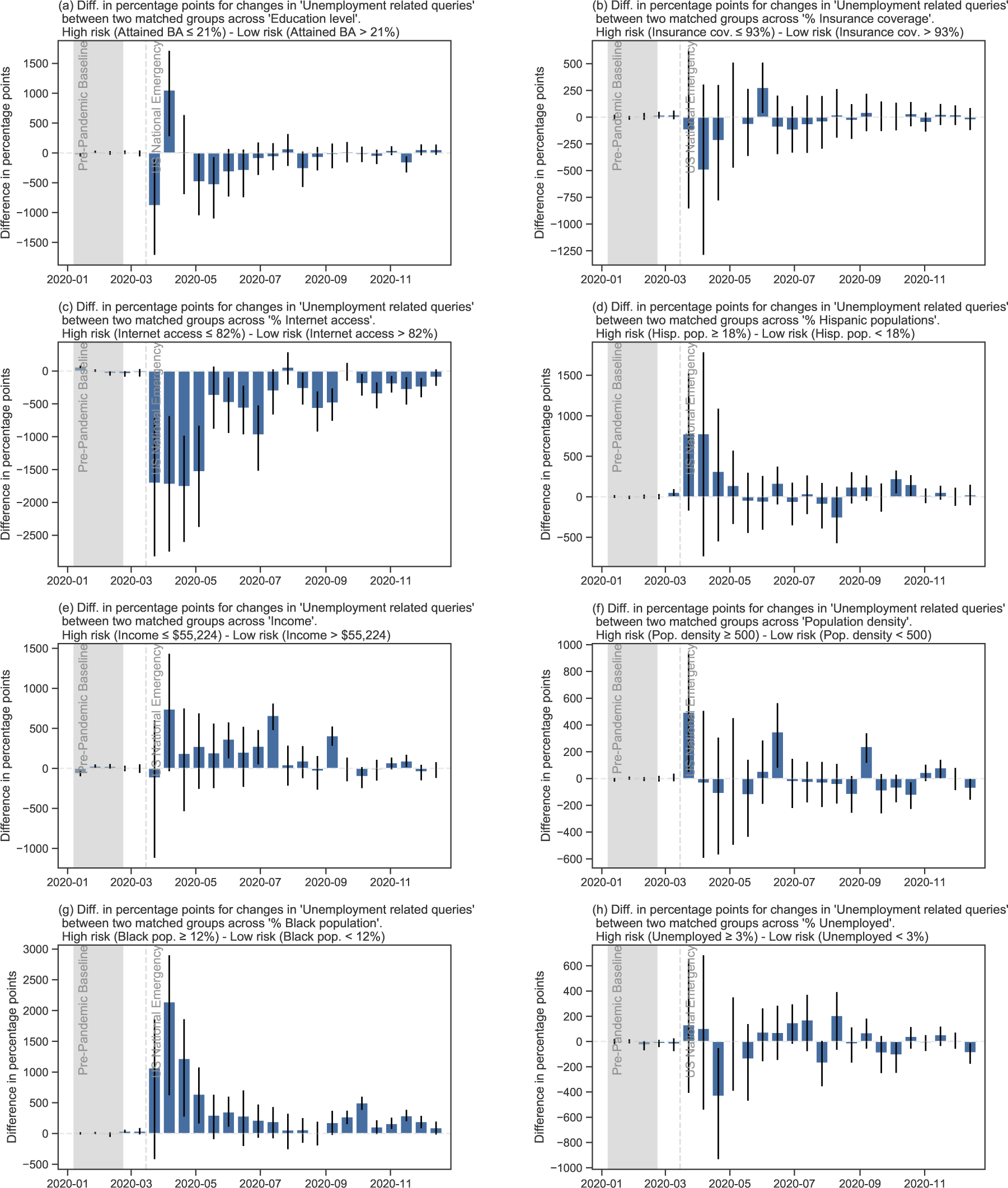
Differences in percentage points for changes in ‘Unemployment related queries’ between two matched groups across eight SDoH factors.

**Figure S16:**
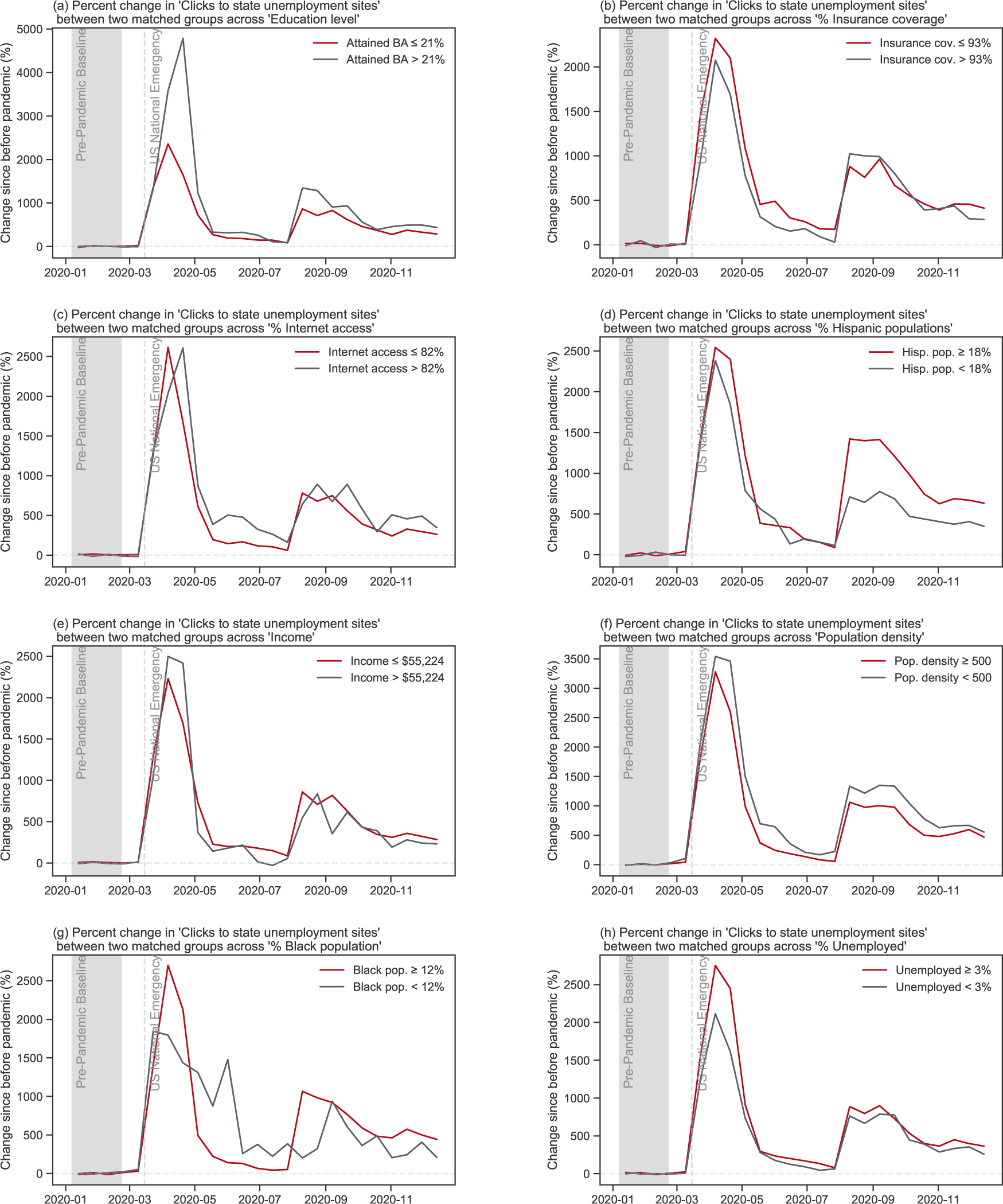
Percent change in ‘Clicks to state unemployment sites’ between two matched groups across eight SDoH factors.

**Figure S17:**
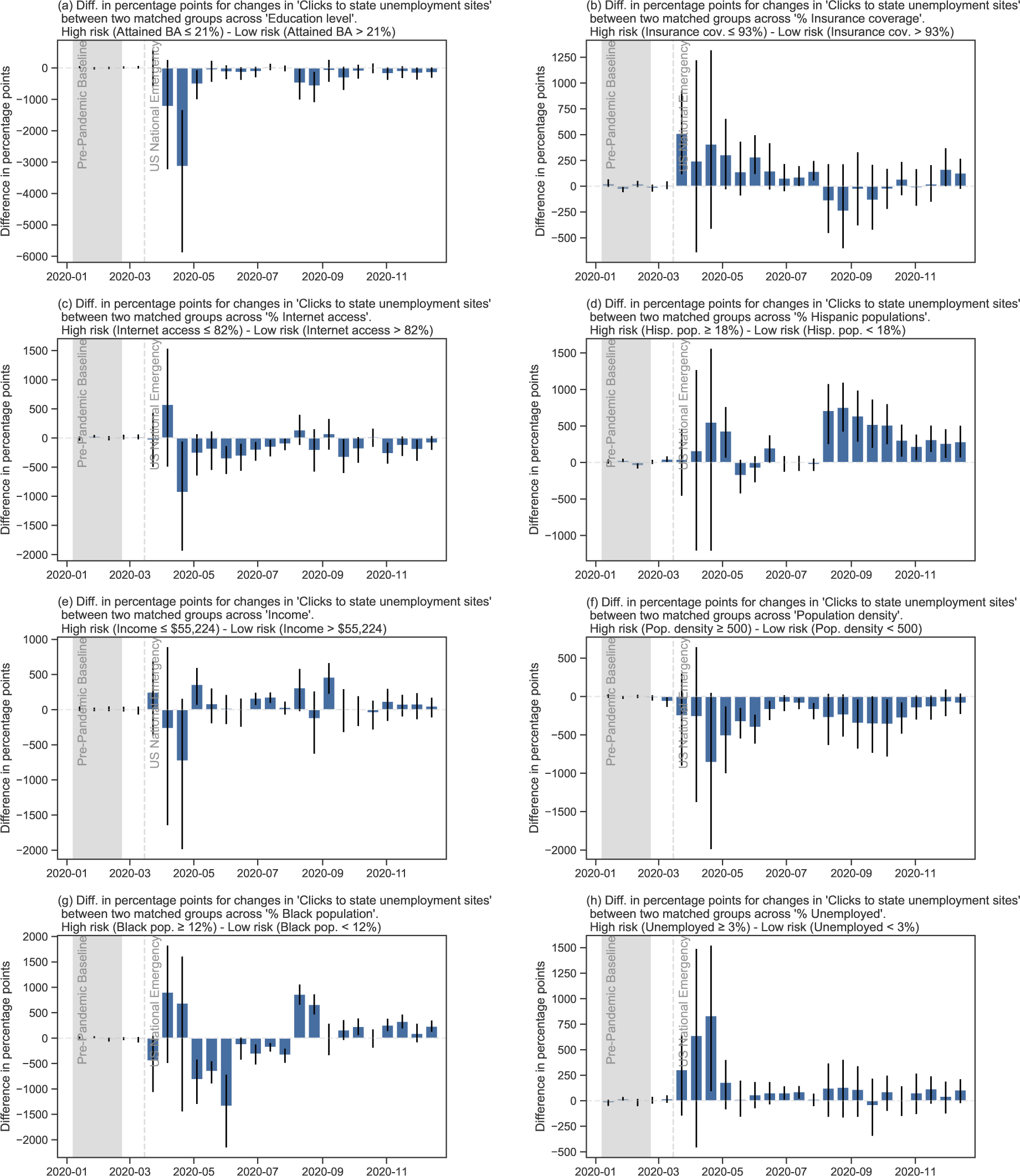
Differences in percentage points for changes in ‘Clicks to state unemployment sites’ between two matched groups across eight SDoH factors.

**Figure S18:**
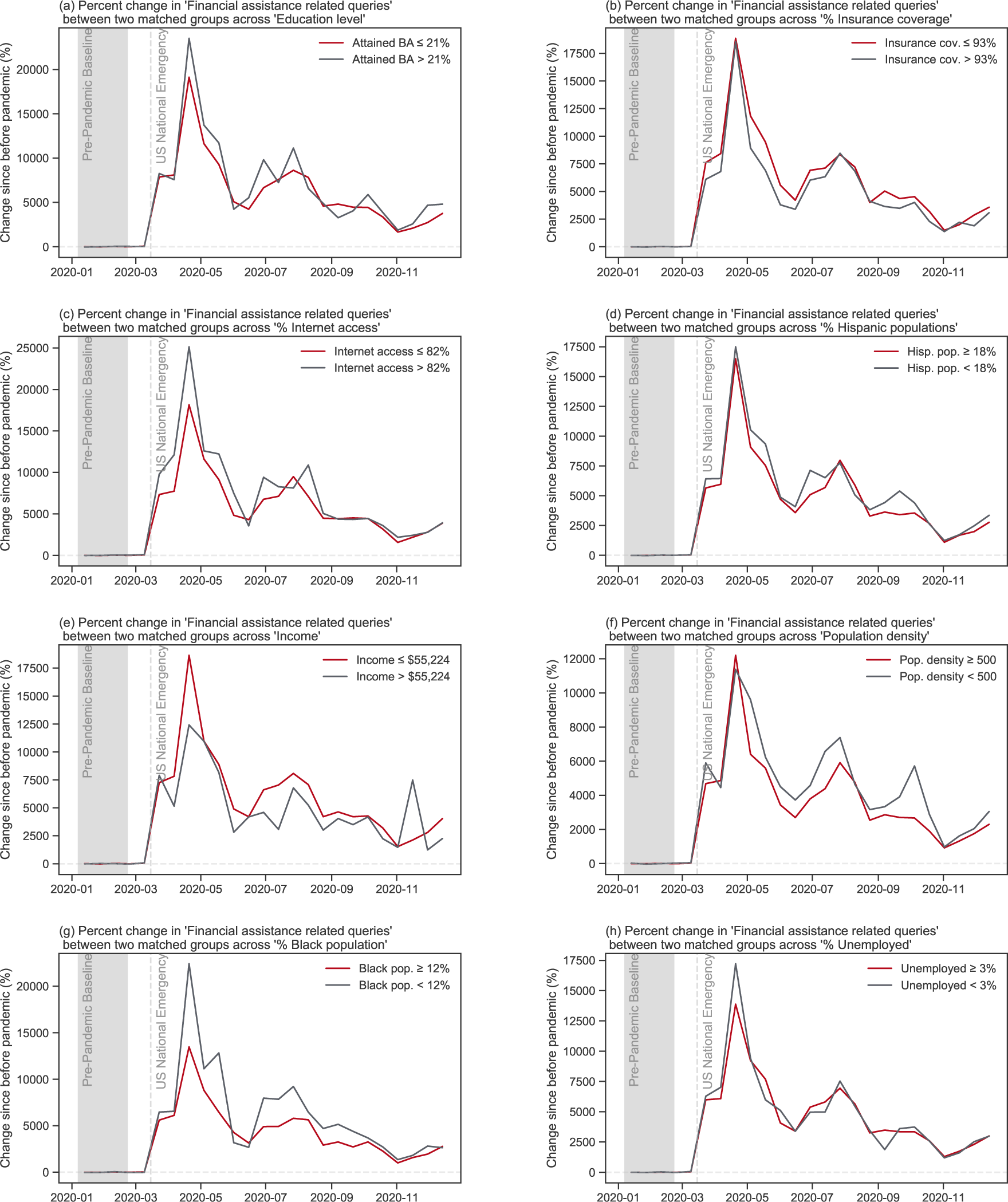
Percent change in ‘Financial assistance related queries’ between two matched groups across eight SDoH factors.

**Figure S19:**
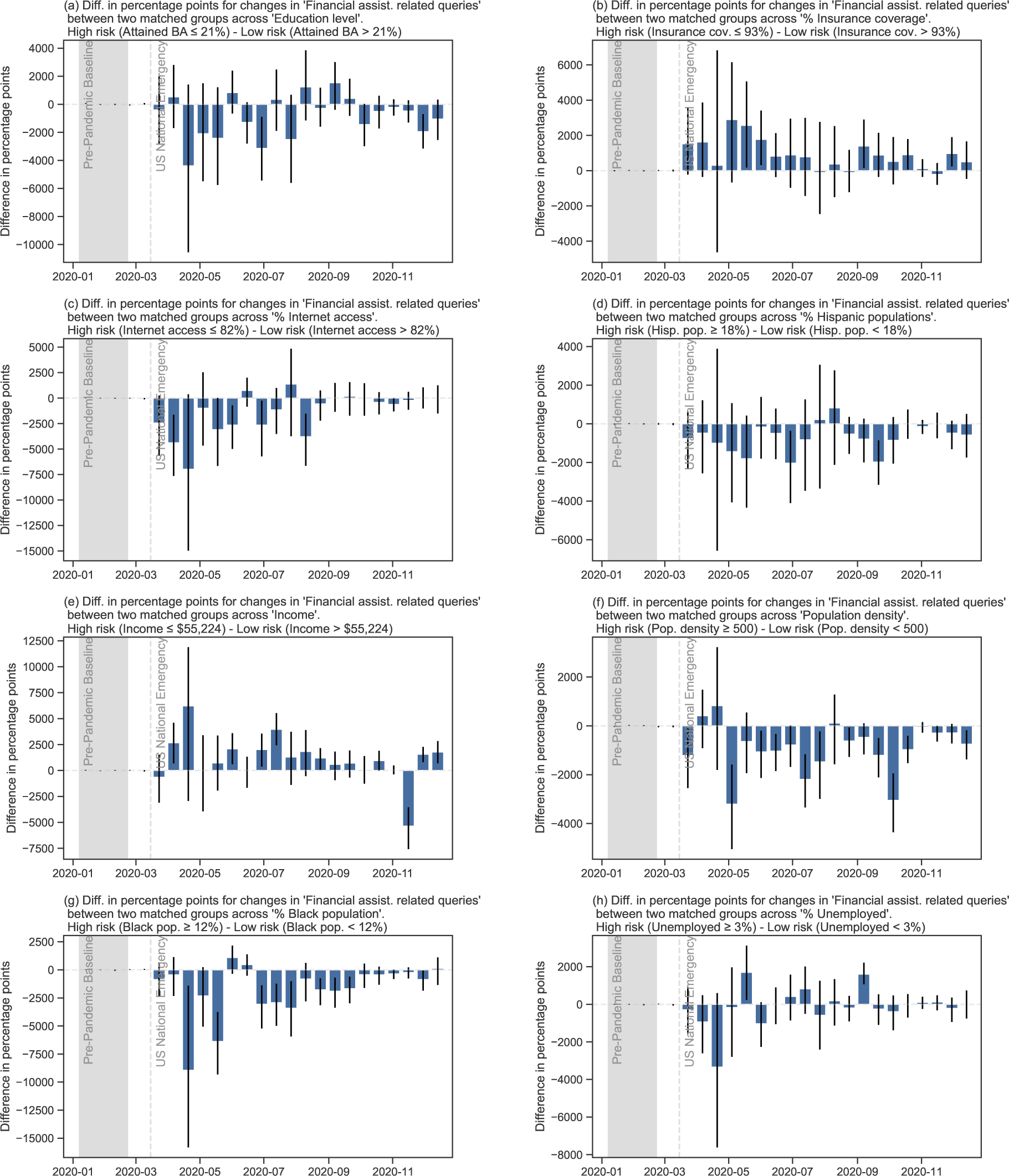
Differences in percentage points for changes in ‘Financial assistance related queries’ between two matched groups across eight SDoH factors.

**Figure S20:**
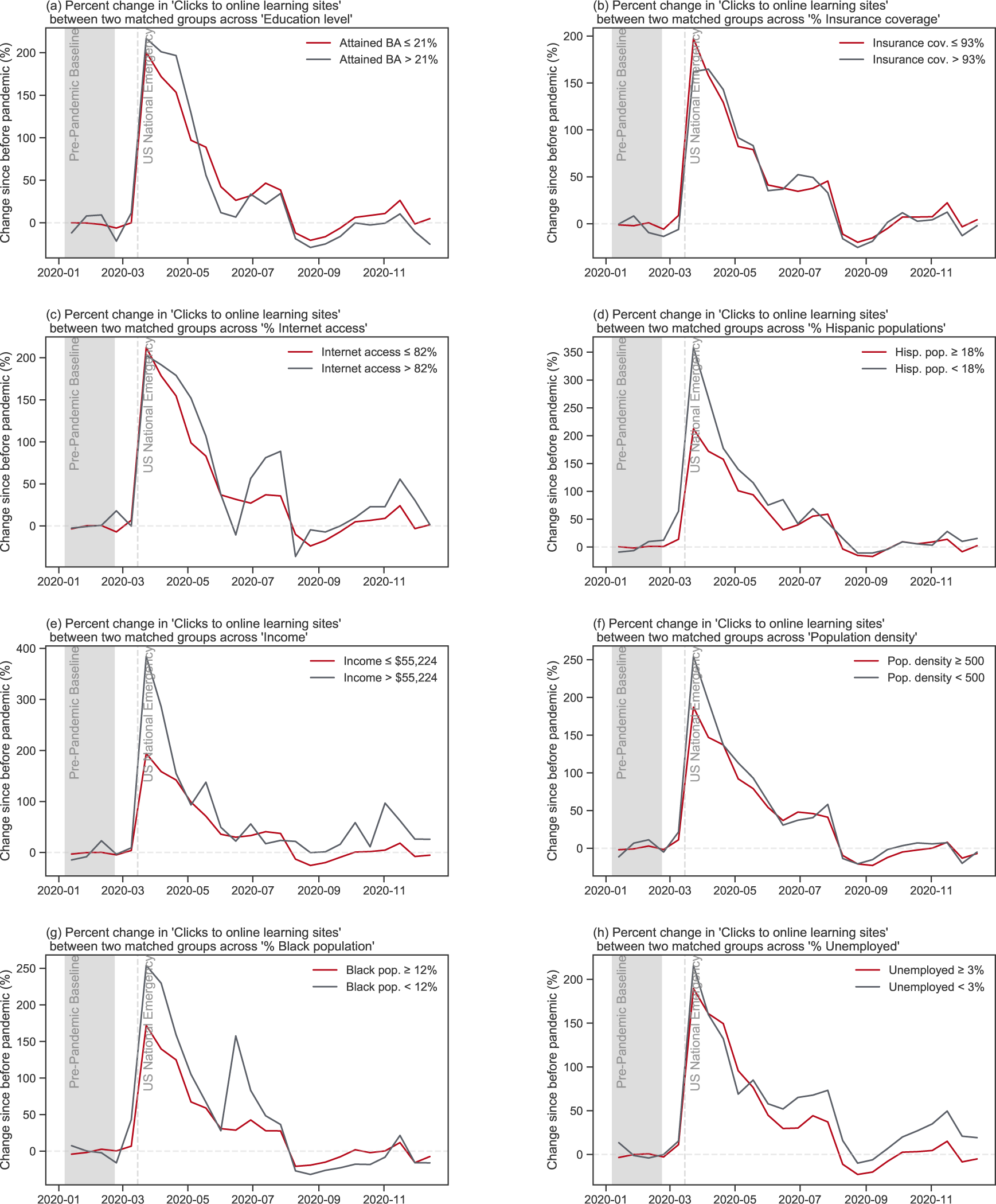
Percent change in ‘Click to online learning sites’ between two matched groups across eight SDoH factors.

**Figure S21:**
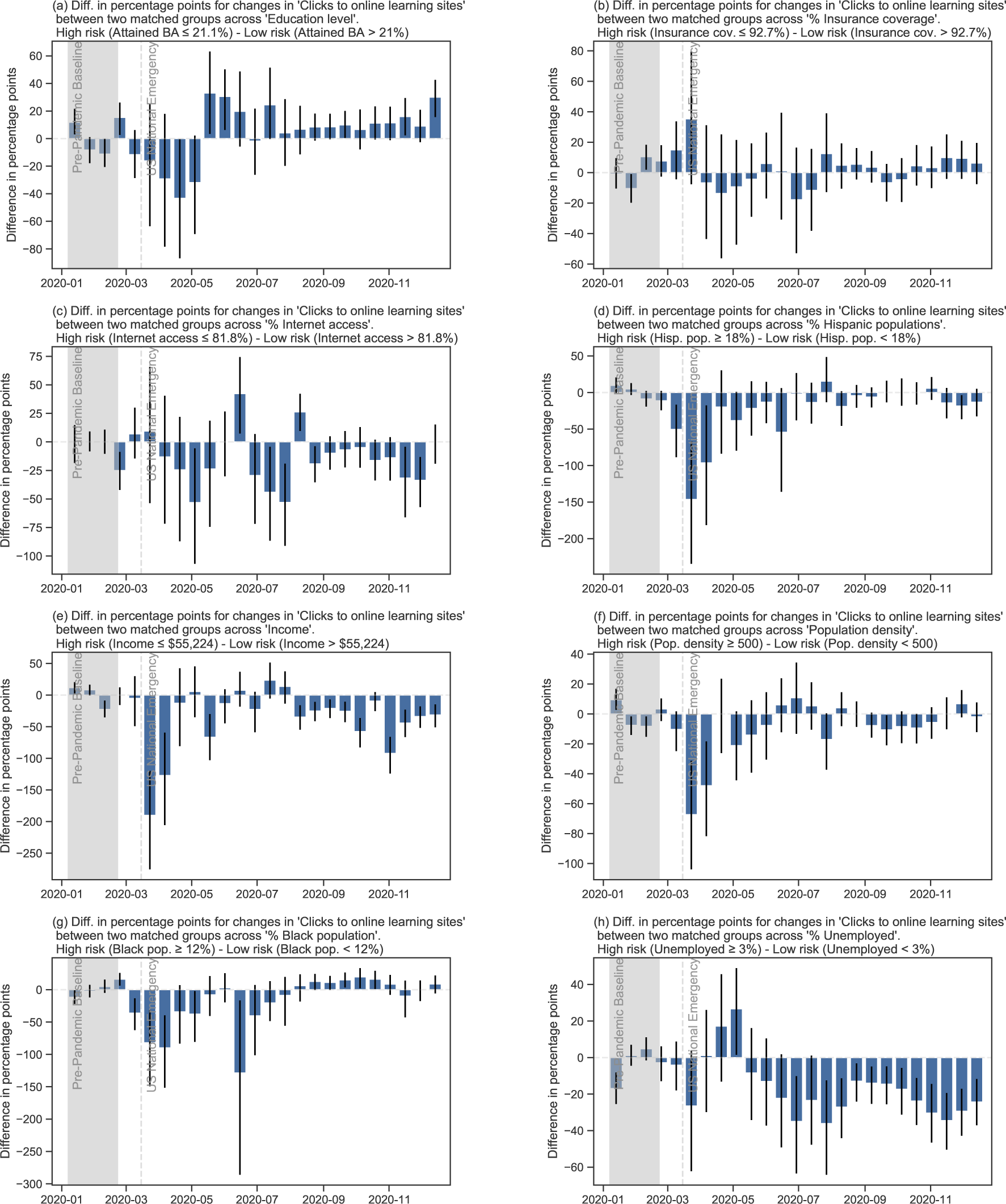
Differences in percentage points for changes in ‘Click to online learning sites’ between two matched groups across eight SDoH factors.

**Figure S22:**
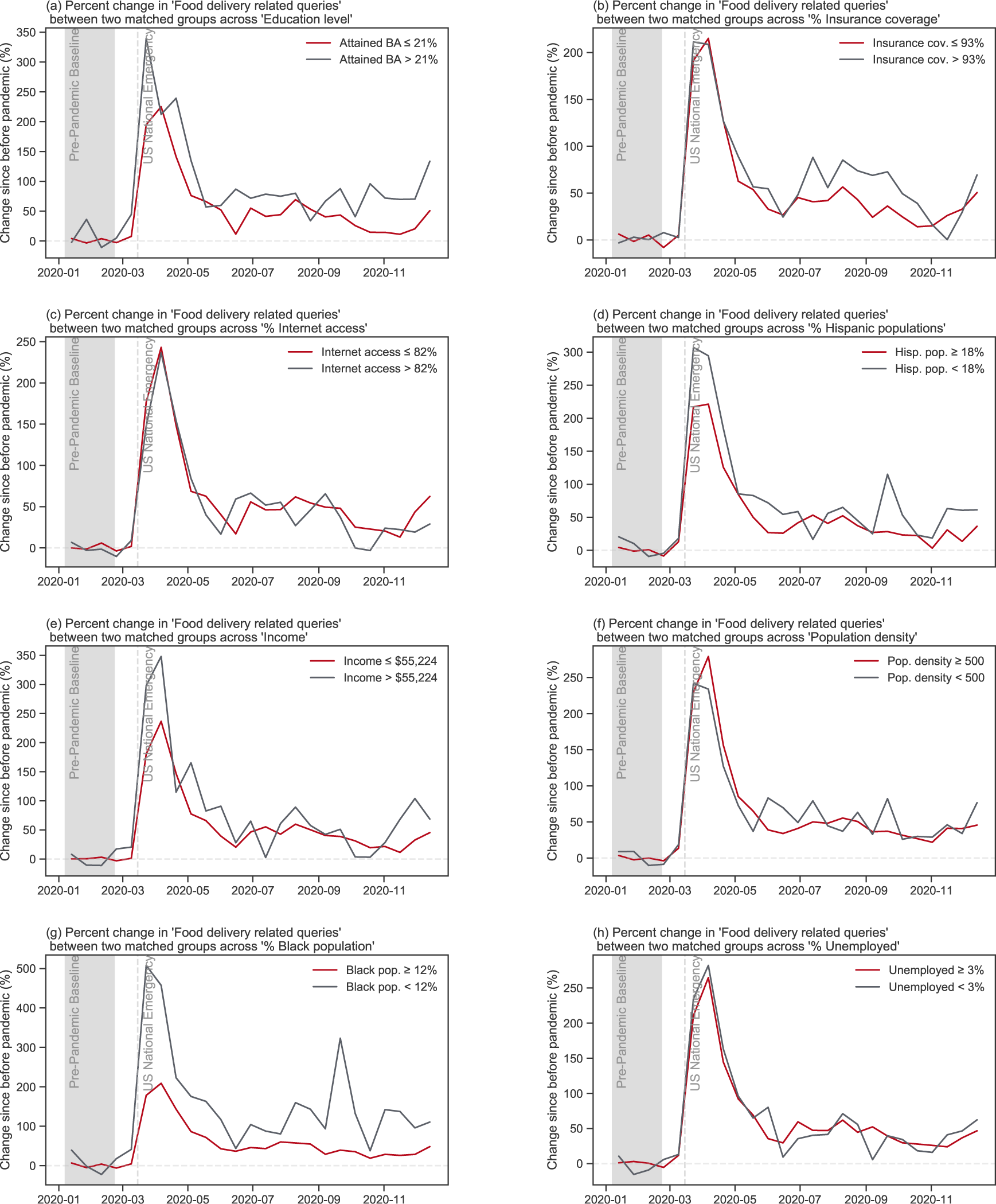
Percent change in ‘Food delivery related queries’ between two matched groups across eight SDoH factors.

**Figure S23:**
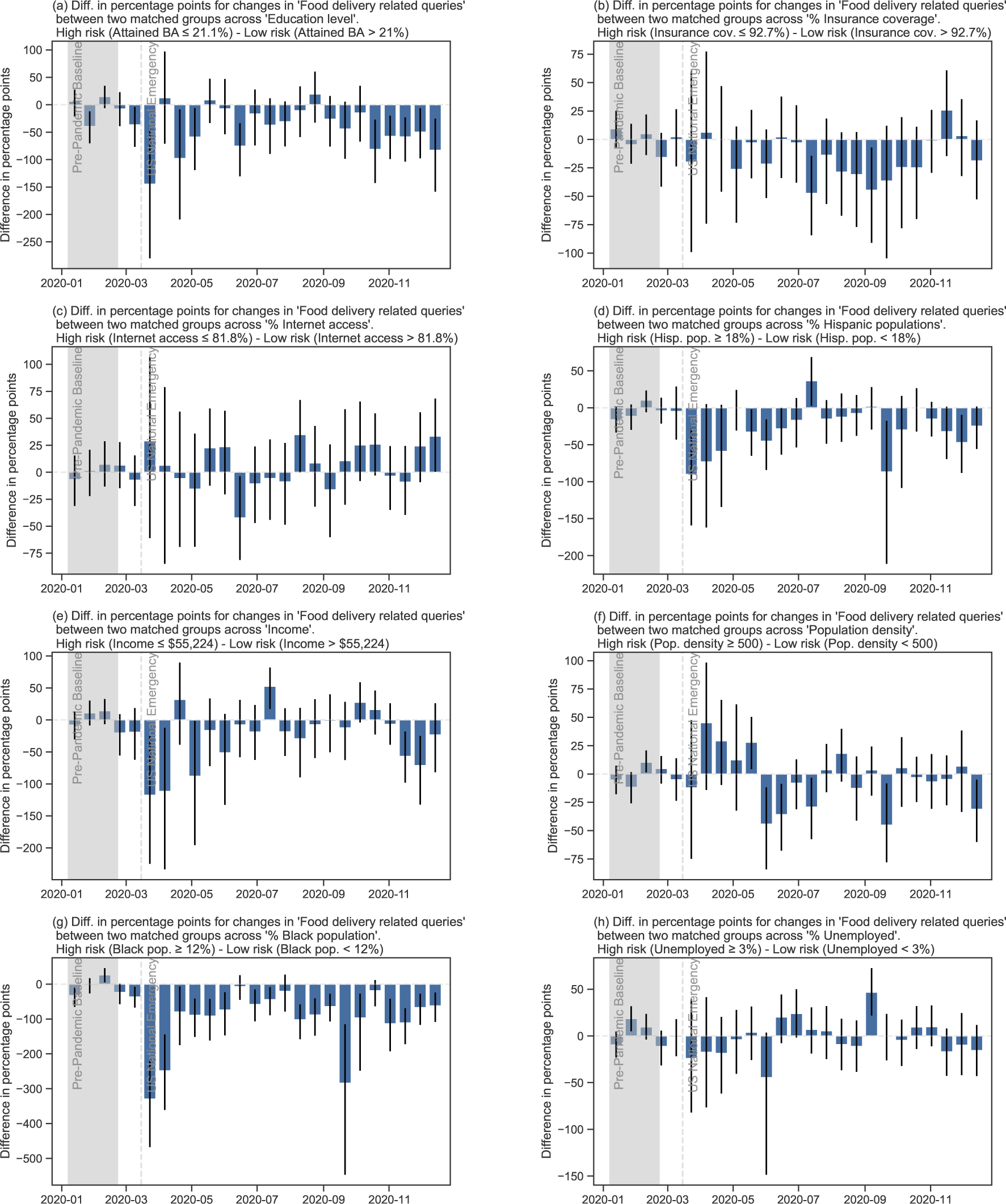
Differences in percentage points for changes in ‘Food delivery related queries’ between two matched groups across eight census variables.

**Figure S24:**
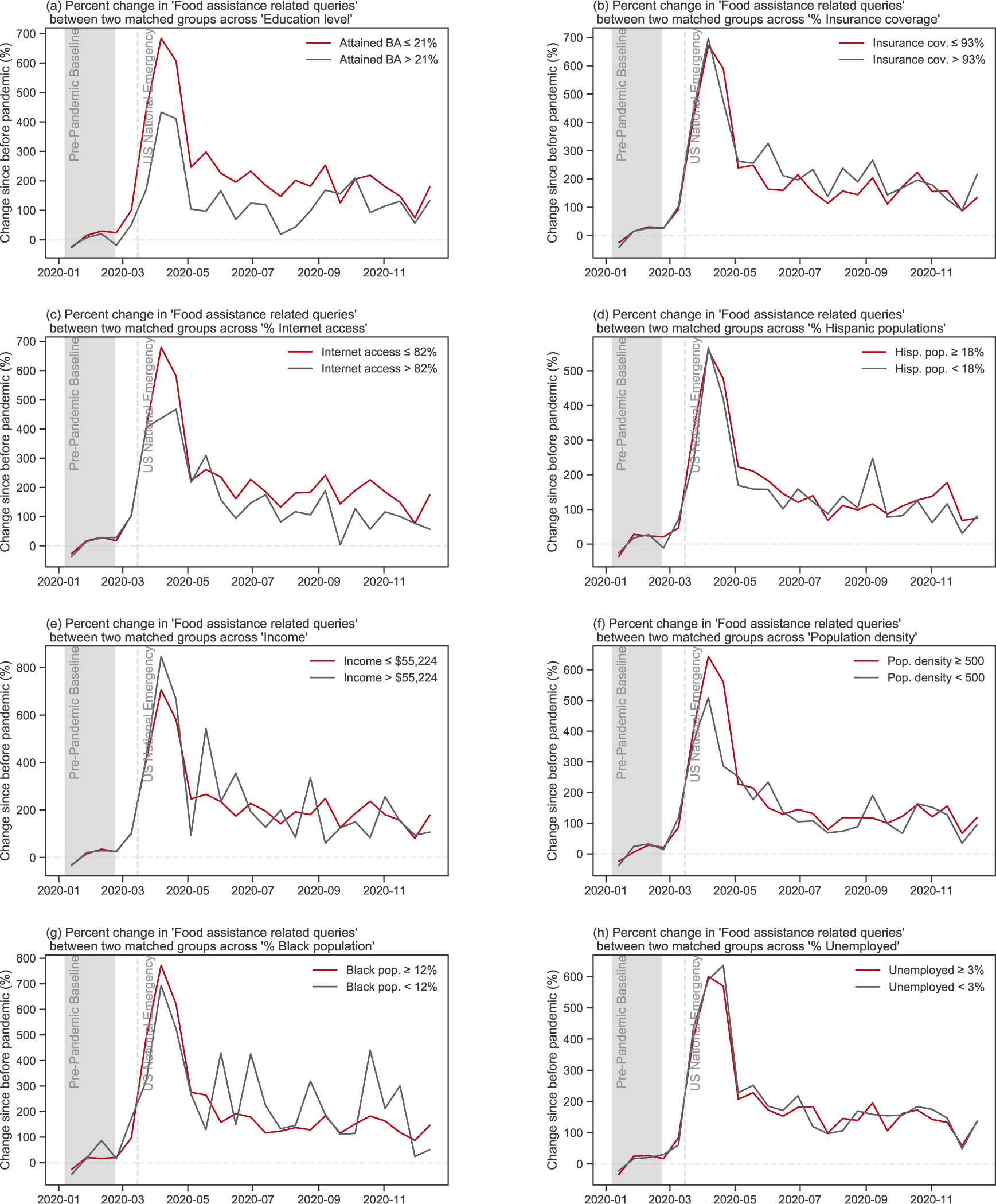
Percent change in ‘Food assistance related queries’ between two matched groups across eight SDoH factors.

**Figure S25:**
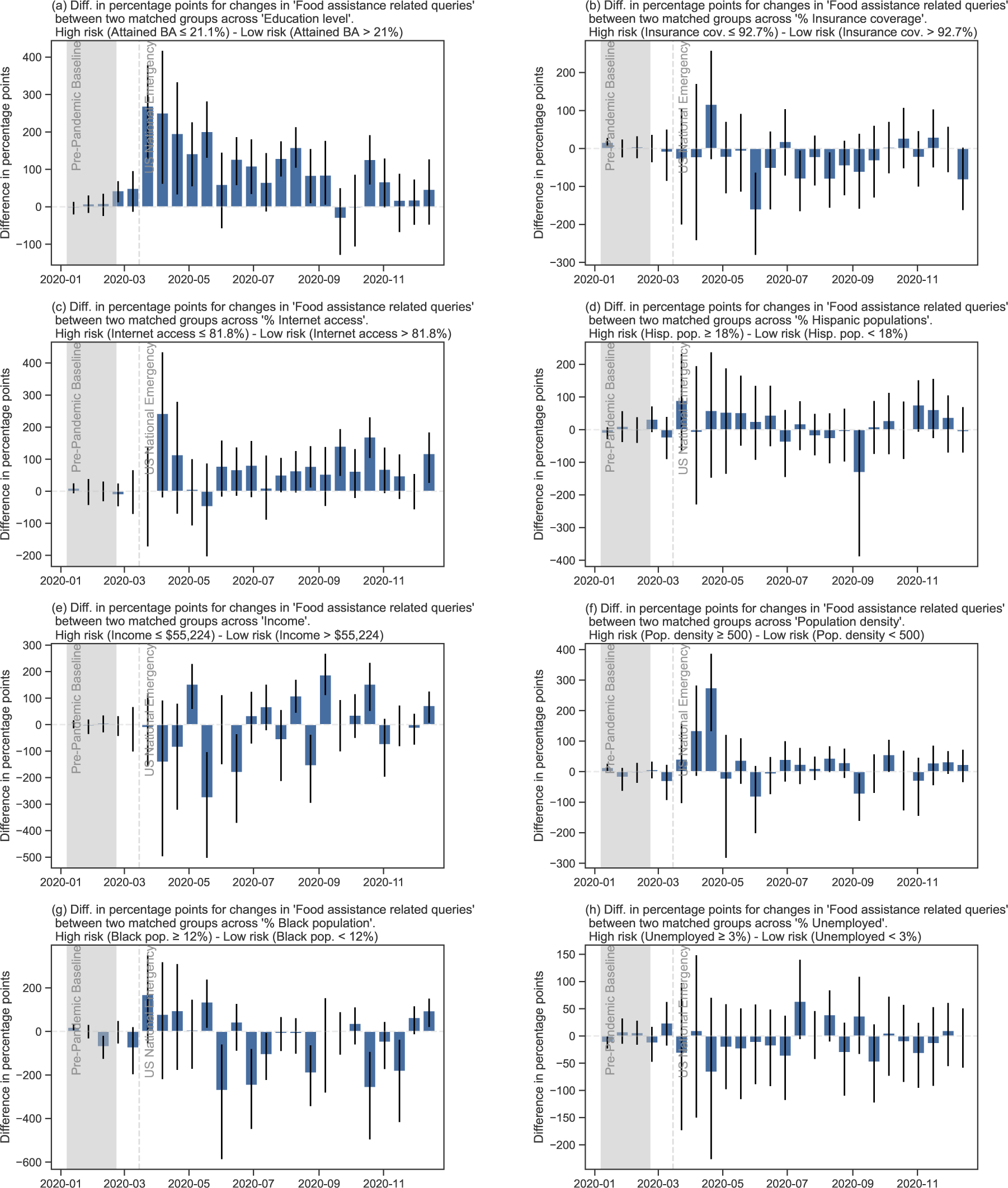
Differences in percentage points for changes in ‘Food assistance related queries’ between two matched groups across eight SDoH factors.

## Footnotes

1 Click share, on the other hand, captures only search-driven traffic to a subset of websites that are instrumented with custom code.

